# LIFT - XAI: Leveraging Important Features in Treatment Effects to Inform Clinical Decision-Making via Explainable AI

**DOI:** 10.1101/2024.09.04.24312866

**Authors:** Mingyu Lu, Chanwoo Kim, Ian Covert, Nathan J. White, Su-In Lee

## Abstract

Clinicians rely on evidence from randomized controlled trials (RCTs) to decide on medical treatments for patients. However, RCTs often lack the granularity needed to inform decisions for individual patients or specific clinical scenarios. Recent advances in machine learning, particularly conditional average treatment effect (CATE) modeling, offer a promising approach for estimating patient-specific treatment effects. Yet, their adoption in clinical practice remains limited because these models often function as “black boxes”, making it difficult to understand which features drive treatment effect heterogeneity among individuals. To overcome these barriers, we introduce LIFT-XAI, a robust and principled framework that interprets ensemble CATE models with Shapley values, thereby accurately identifying unique features and patient subgroups driving the treatment effects. We validate LIFT-XAI on real-world clinical data from four RCTs comprising over 42,000 patients. We demonstrate that LIFT-XAI uncovered fasting glucose as the primary factor explaining conflicting outcomes between two major blood pressure trials (SPRINT and ACCORD) and discovered age as a critical treatment modifier in a new clinical setting–a finding later validated by a subsequent RCT. By enabling such powerful cross-cohort analysis and the discovery of novel patient subgroups, LIFT-XAI advances precision medicine across diverse settings, from chronic disease management to acute care.

## Introduction

Assessing the effectiveness of an intervention is a core challenge across many high-stakes domains ^1,2^, especially in medicine. Healthcare professionals use available evidence to determine which treatments are most likely to improve health outcomes for individual patients ^2^. In this context, randomized controlled trials (RCTs) are considered as the gold standard ^3^ for establishing the existence of a significant treatment effect. Specifically, RCTs estimate the average treatment effect (ATE) of an intervention when applied to cohort of patients pre-selected by several criteria (Figure 1a). Building on this foundation, efforts to understand why treatments differ across subpopulations have traditionally relied on secondary subgroup analyses of RCTs, which stratify patients based on predefined covariates within a single study cohort ^4^. However, such analyses offer limited insight, as they are often underpowered to detect individual-level differences ^2,5^ (Figure 1b).

**Fig. 1.**
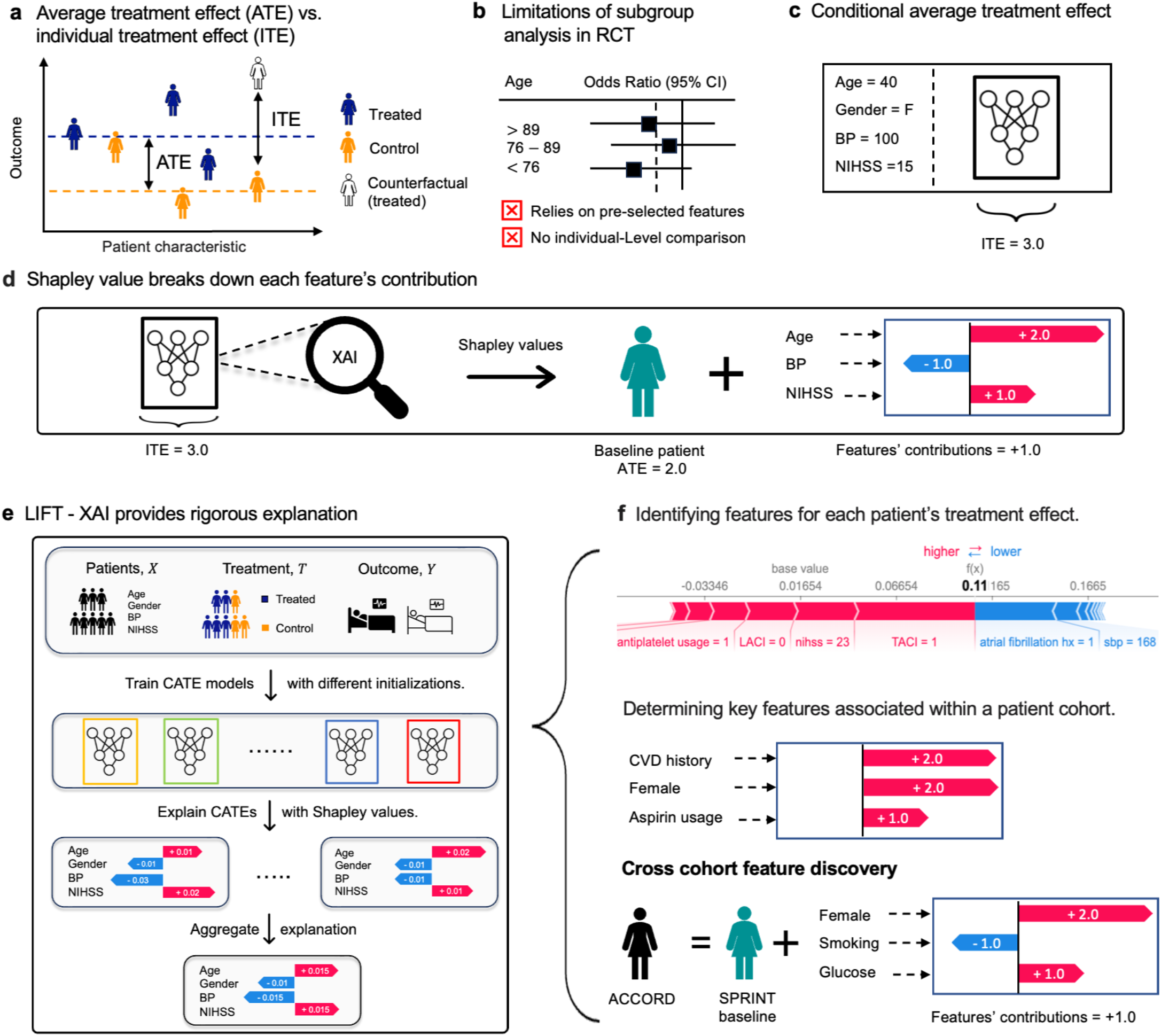
Conceptual background and the LIFT-XAI framework. (a) The average treatment effect (ATE) measures the mean difference in outcomes between treated and control groups, while the individual treatment effect (ITE) captures the effect for a single patient relative to their “counterfactual” outcome (e.g., their outcome if untreated versus if treated). (b) Conventional subgroup analyses stratify patients based on predefined categorical features but do not allow for patient-level comparisons. (c) Conditional average treatment effects (CATEs) capture heterogeneity by conditioning on patient covariates. (d) Explainable AI (XAI) enables explaining black-box CATE models; Shapley values quantify feature contributions relative to a baseline (add some description about the baseline patient) patient. b(e) **LIFT-XAI:** proposed framework that explains ensemble CATE models with Shapley values, aggregating stable attributions across CATE models to reveal robust, clinically meaningful signals that support decision-making and exploratory analysis. (f) Use cases for LIFT-XAI. Top: Individual-level feature contributions derived from LIFTXAI for a representative patient. Middle: LIFT-XAI provides cohort-level feature attribution of treatment effects. Bottom: LIFT-XAI enables cross-cohort comparison, identifying features that drive differences in treatment effects between similar patient populations.

These limitations highlight the need for approaches that move beyond population estimates and provide insights into how treatment responses differ among individuals. Recent work has explored machine learning–based methods such as conditional average treatment effect (CATE) modeling ^6^, which estimate individual treatment effects (ITE) as a function of patient covariates (e.g., age, sex, health status) (Figure 1c). In particular, neural network–based models show strong potential for capturing complex, nonlinear treatment heterogeneity^7–11^. However, their clinical utility remains limited. Effective clinical decision-making requires understanding how individual patient characteristics influence treatment effects, yet we often lack insight into how these “black-box” models use such information. Bridging this interpretability gap is crucial, as it could not only foster broader clinical adoption of CATE modeling but, more importantly, help elucidate an important question–*why do individual responses to treatments vary?* Furthermore, to better support clinical decision making, it is essential to ask *whether individual-level insights can be elevated to the cohort or subgroup level?* Aligning such generalized explanations with established clinical discovery can strengthen clinicians’ confidence in CATE model interpretations and promote their integration into real-world clinical workflows.

Answering these questions could advance our understanding of individual treatment effect heterogeneity and better support clinicians in decision-making.

To this end, we propose *LIFT-XAI* (**L**everaging **I**mportant **F**eatures in **T**reatment effects to inform clinical decision-making via e**X**plainable **AI**), a framework to improve the interpratability of CATE models through model ensembling and Shapley values (Figure 1d-e). Our approach presents several breakthroughs over existing approaches for using *explainable AI (XAI)* in CATE modeling. First, whereas prior methods that only provide population-level feature importance ^12–14^, LIFT-XAI provides fine-grained, individual-level attributions based on Shapley values ^15,16^. By comparing a given patient to a “baseline” patient, Shapley values decompose the treatment effect into feature contributions, thereby revealing what feature drives treatment effects. Second, LIFT-XAI integrates model ensembling with Shapley value to enhance reliability and reduce variance. In contrast, most existing approaches rely on explanations from a single model ^17^, which can be unstable due to the stochastic nature of training, such as random initialization or hyperparameter variability ^18^. Third, by aggregating these reliable local explanations, LIFT-XAI also recovers global-level insights, thereby bridging the gap between personalized and population-level interpretability (Figure 1f). To summarize, through explaining ensemble CATE using Shapley values, LIFT-XAI delivers reliable attribution scores, offering a robust framework for understanding heterogeneous treatment effects.

To facilitate clinical translation, moving beyond prior studies that evaluated only semi-synthetic data or small clinical cohorts ^19,20^, we benchmarked LIFT-XAI against existing explanation methods using four large real-world RCTs comprising over 42,000 patients. We found that LIFT-XAI produces stable and rigorous individual-level explanations while accurately identifying features that are important at the population level. Notably, the features identified by LIFT-XAI closely aligned with key factors reported in the aforementioned clinical studies. Furthermore, we tested LIFT-XAI against the two most common hurdles present when applying RCTs to real-world practice–differences in patient characteristics and alternative clinical practice settings–by introducing a *cross-cohort attribution analysis*. By comparing patients within one cohort to baseline patients across cohorts, we identify and quantitatively validate influential features that explain the divergent outcomes of the ACCORD and SPRINT trials ^21,22^. Moreover, using in-hospital trauma patients as a baseline, we identify key features associated with pre-hospital tranexamic acid (TXA) administration, recapitulating findings from an independent RCT. This demonstrates the potential of LIFT-XAI to uncover clinically meaningful features across settings without the need for costly RCTs.

## Results

LIFT-XAI is a framework that delivers rigorous explanations at both the global and individual levels by integrating ensemble CATE modeling with Shapley value. Before demonstrating how LIFT-XAI reveals clinically meaningful features underlying treatment heterogeneity, we evaluate these two key components to establish its effectiveness.

### Ensemble CATE outperforms a single model

Building on prior evidence that model performance strongly influences explanation quality ^19^, we propose ensemble strategies to obtain high-performing CATE estimators. To evaluate the effectiveness of our proposal, we first trained and compared multiple CATE models of different types, including Doubly Robust (DR)-Learner ^10^, regression-adjustment(RA)-learner ^23^, and self-interpretable models such as Causal Forest and Linear DR (see Methods). We evaluated their performance across four large clinical trial datasets, including IST-3, CRASH-2, SPRINT, and ACCORD ^21,22,24,25^. Model performance was assessed on prediction error metrics (e.g., pseudo-outcome surrogates) and incremental gain metric (e.g., Qini and uplift scores), all calculated on held-out test sets (see Section B.3).

Our results show that the DR-Learner outperformed other learners on IST-3, CRASH-2, and SPRINT, with the lowest pseudo–outcome errors (0.921, 0.684, and 0.473) and highest Qini scores (0.217, 0.072, and 0.066), respectively (Table S.2). In contrast, the RA-Learner achieved the best performance on ACCORD. Self-interpretable models such as Causal Forest and Linear DR underperformed relative to neural network-based learners.

Furthermore, building ensembles from the best-performing learner consistently outperforms single models (Table 1). In addition, when we computed the cohort-level average treatment effects (ATEs) based on ensemble CATEs, we found that they aligned well with reported trial results for IST-3 and CRASH-2. Although estimates for SPRINT and ACCORD were slightly higher, they remained more accurate than those from single models (Table S.3).

**Table 1.** Model performance on IST-3, CRASH-2, SPRINT, and ACCORD. Metrics include pseudo-surrogate outcomes ( ℰ_if-pehe_, ℰ_R_, ℰ_DR_) and incremental gain scores (Qini and Uplift). Results are reported as mean *±* standard deviation across random seeds. Higher values indicate better performance for Qini and Uplift, whereas lower values indicate better performance for ℰ_R_, and ℰ_DR_.

|  | IST-3<sup>24</sup> |  | CRASH-2<sup>25</sup> |  | SPRINT<sup>22</sup> |  | ACCORD<sup>21</sup> |  |
| --- | --- | --- | --- | --- | --- | --- | --- | --- |
|  | DR-Learner | DR-Learner (Ens.) | DR-Learner | DR-Learner (Ens.) | DR-Learner | DR-Learner (Ens.) | RA-Learner | RA-Learner (Ens.) |
| Qini | 0.284 (0.034) | **0.358 (0.016)** | 0.136 (0.004) | **0.186 (0.004)** | 0.066 (0.003) | **0.117 (0.003)** | 0.096 (0.007) | **0.212 (0.008)** |
| Uplift | 0.300 (0.025) | **0.395 (0.019)** | 0.248 (0.005) | **0.316 (0.008)** | 0.125 (0.005) | **0.212 (0.005)** | 0.183 (0.012) | **0.358 (0.011)** |
| $\mathcal{E}_R$ | 0.521 (0.008) | **0.520 (0.008)** | **0.373 (0.003)** | 0.373 (0.003) | 0.241 (0.004) | **0.236 (0.006)** | 0.305 (0.005) | **0.300 (0.005)** |
| $\mathcal{E}_{DR}$ | 0.921 (0.016) | **0.919 (0.015)** | 0.684 (0.005) | **0.678 (0.004)** | **0.473 (0.007)** | 0.472 (0.010) | 0.592 (0.009) | **0.580 (0.009)** |

Together, these results demonstrate that ensemble CATE learners yield reliable treatment effect estimates consistent with real-world trial findings, making them the most suitable explainees. Accordingly, in the following, we select the best-performing model type and use its ensemble as the explainee within the LIFT-XAI framework.

### Shapley value provides rigorous explanation in real-world clinical data

We next benchmarked our chosen explanation method, Shapley value^15,16^, against alternative approaches, against other explanation approaches by assessing their local fidelity, global sufficiency, and robustness (see Methods) to show the advantage of our choice.

#### Local fidelity

We evaluated whether features ranked as important by each method meaningfully affected predictions using insertion–deletion tests ^26^. Figure 2 (a) shows the deletion and insertion curves for the SPRINT dataset. In the deletion test (Figure 2 (a-top), pseudo-surrogate error increased as top contributing features were removed, with Shapley values outperforming Integrated Gradients (IG), LIME, and Saliency. In the insertion test (Figure 2 (a-bottom), IG performed comparably to Shapley values. Overall, when comparing Area Under the Curve (AUC), our results show that the Shapley value method achieves the highest normalized AUC in the ACCORD, SPRINT, and CRASH-2 datasets, Section D.3. In contrast, for the IST-3 dataset, LIME performs best in insertion, while the Shapley value performs best in deletion. These results indicate that Shapley explanations faithfully capture the features driving individual-level treatment effect heterogeneity.

**Fig. 2.**
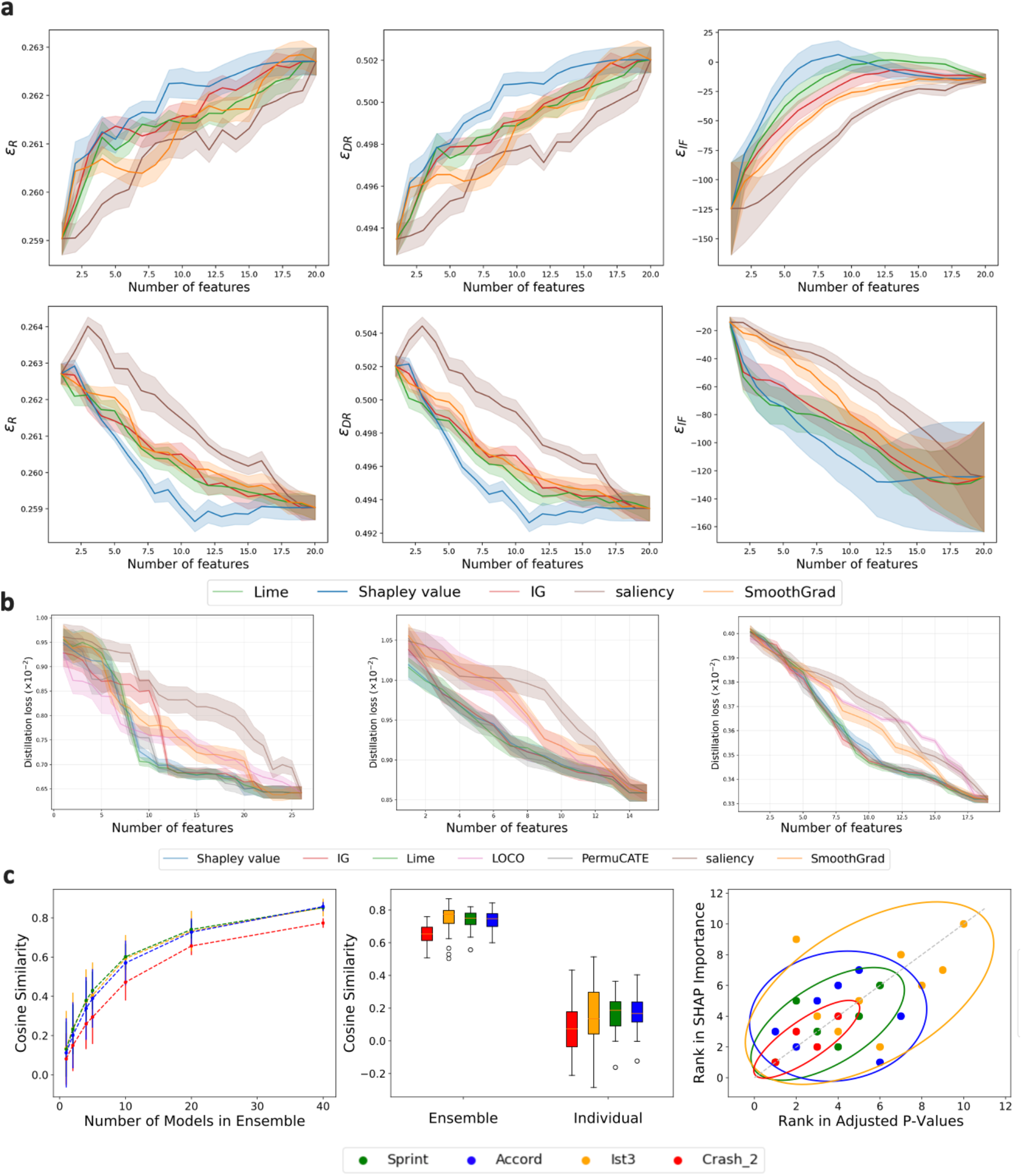
Evaluation of explanation methods for heterogeneous treatment effect models. **(a)** Local fidelity: deletion (top) and insertion (bottom) experiments on the SPRINT dataset using pseudo-outcome–based metrics. The *x*-axis denotes the number of features removed or included, and the *y*-axis reports surrogate error. (left to right). **(b)** Global sufficiency: Knowledge distillation performance across datasets, ACCORD, IST-3, and CRASH-2 (left to right). The *x*-axis denotes the number of features retained in student models, and the *y*-axis shows the distillation loss (MSE) relative to the ensemble teacher. **(c)** Consistency evaluation of explanation methods. (Left) Cosine similarity of feature attributions as a function of the number of models in the ensemble. (Middle) Distribution of cosine similarities for ensemble-averaged versus individual model explanations. (Right) Concordance between Shapley importance ranks and interaction *p*-value ranks across clinical datasets, with 95% confidence ellipses.

#### Global sufficiency

We then examined global (cohort-level) feature importance using our proposed distillation benchmark (Methods), which measures whether student models with top-ranked features can reconstruct ensemble outputs. For local attribution methods, features were ranked by their average absolute attribution across individuals. In ACCORD, Shapley values achieved the lowest distillation loss under restricted feature budgets, highlighting their efficiency in capturing features most relevant for CATE estimation, Figure 2 (b). In SPRINT and IST-3, Shapley value and LIME perform similarly to global feature importance approaches such as PermuCATE^13^ and outperform LOCO ^12^. We also qualitatively examined the top features identified by each method (Table S.8). In CRASH-2, both IG and Shapley values highlighted clinically meaningful factors such as GCS score and injury type, whereas Saliency tended to emphasize less informative features, such as capillary refill time.

#### Consistency

Finally, we evaluated the stability of Shapley explanations by comparing global attributions from single versus ensemble models using cosine similarity. Single-model explanation exhibited low similarity and high variance, with scores of 0.13, 0.15, 0.15, and 0.21, Figure 2 (c-middle). By contrast, ensemble-based Shapley explanations were far more consistent: the average similarity increased from 0.6 with 10 models to 0.8 with 20 models (Figure 2 c-left), highlighting the enhanced reliability and stability.

Taken together, these findings show that Shapley values yield high-fidelity explanations at both local and global levels, while ensembling further enhances robustness. This motivates our proposed **LIFT-XAI**, which combines ensemble CATEs with Shapley values to provide reliable feature attributions for treatment effect estimation.

### Validating LIFT-XAI for Cohort-Level Feature Discovery

We next evaluated whether LIFT-XAI could recover known treatment effect modifiers at the cohort level. Using well-established RCTs with known influential features, we assessed whether LIFT-XAI could rediscover these factors, thereby validating its clinical utility. To do this, we compared its global feature rankings with those reported in the original studies using Spearman’s rank correlation ^27^. For these studies, reported interaction p-values were used as proxies for feature ranking ^28^. A lower interaction p-value indicates a higher likelihood of a feature being a treatment effect modifier. Our findings show a significant correlation between Shapley rankings and reported features, with values of 0.8, 0.54, and 0.6 for the CRASH-2, IST-3, and SPRINT, respectively, Table 2; Figure 2 (c-right). In contrast, the ACCORD study shows a low correlation (0.05), which is expected, as no significant features were reported ^21^.

**Table 2.** Correlation between ranks based on LIFT-XAI and interaction p-values across RCTs.

| Dataset | Correlation (Corr) | p-value | Number of Reported Features |
| --- | --- | --- | --- |
| CRASH-2 | 0.80 | 0.11 | 4 |
| IST-3 | 0.54 | 0.09 | 10 |
| SPRINT | 0.60 | 0.12 | 6 |
| ACCORD | 0.05 | 0.90 | 7 |

In the following sections, we will demonstrate how LIFT-XAI can be used to analyze clinical trial data. In what follows, we use the terms Shapley values and LIFT-XAI interchangeably.

### Use cases 1: Decomposing rt-PA treatment effect into features’ contribution with LIFT-XAI

We obtained individual explanations through LIFT-XAI for patients withing IST-3 trials that assesses the efficacy of intravenous rt-PA in acute ischaemic stroke patients. Unlike traditional subgroup analysis, which requires dividing patients into subgroups and calculating risk or odds ratios, LIFT-XAI allows direct analysis of feature impact at both individual and cohort levels. It provides individual-level explanations ^16,29^ by breaking down the total treatment effect into contributions from each feature for every patient (Figure 3a).

**Fig. 3.**
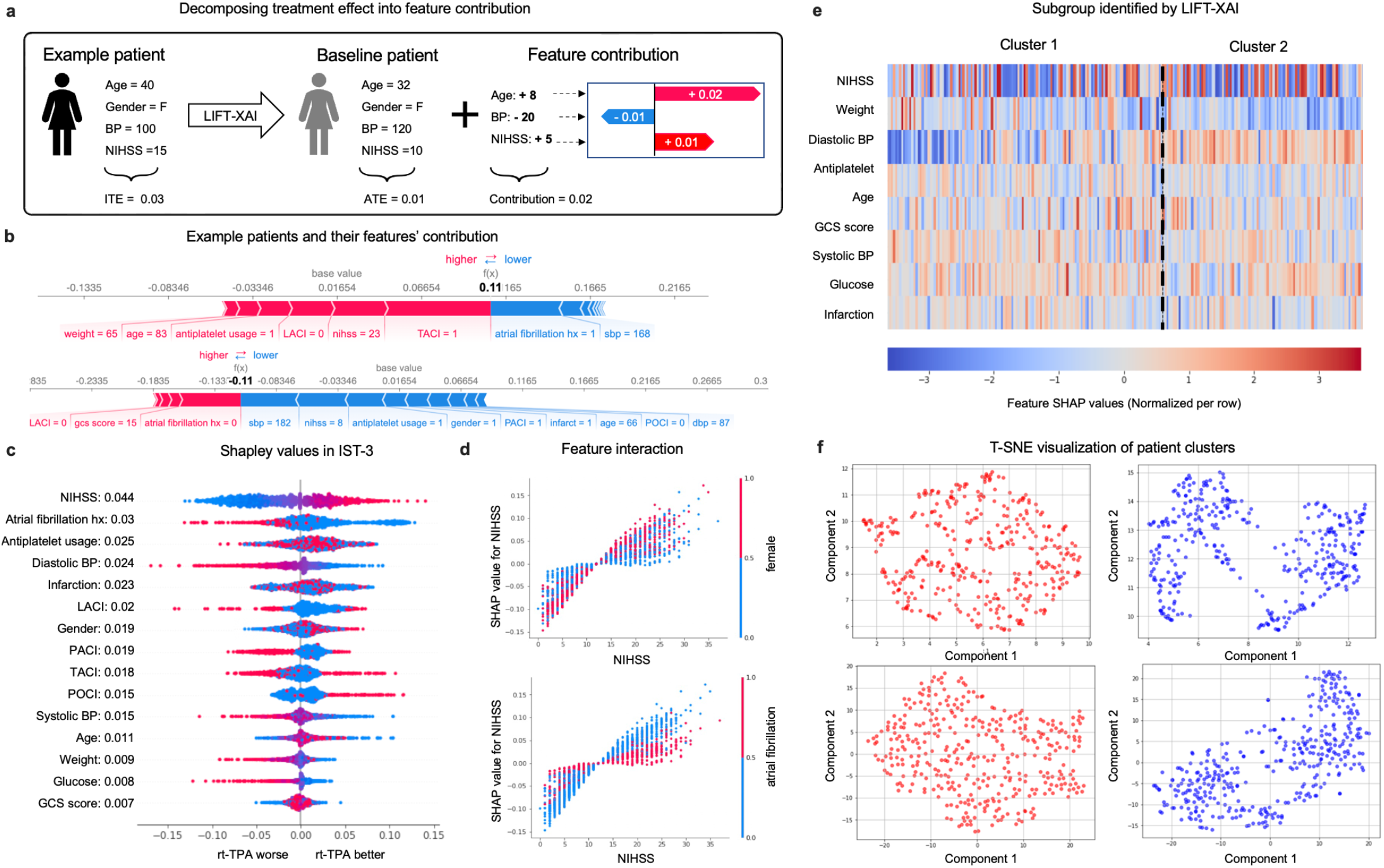
Analyzing the IST-3 Study with LIFT-XAI: (a) LIFT-XAI decomposes feature contributions for an example patient by comparing them with a baseline patient whose feature values correspond to the average across the IST-3 trial. (b) Shapley values for example individuals, where red indicates positive attributions and blue represents negative attributions. (c) IST-3 summary plot showing features on the y-axis sorted by mean absolute Shapley values and on the x-axis by their corresponding Shapley values. Colors indicate feature values, with red for higher and blue for lower. (d) Combined Shapley values (left y-axis) and feature values pairs (x-axis and right y-axis) of NIHSS with gender (top) and atrial fibrillation (bottom). For binary features, the red dot indicates a feature value of 1, while blue indicates 0. (e) Hierarchical clustering of IST-3 patients with NIHSS scores between 6 and 14 based on their corresponding Shapley values. (f) Hierarchical clustering based on Shapley value and corresponding heatmap. t-SNE visualizations of patient clusters.

In Figure 3b, the upper plot shows an example patient’s feature contribution. This patient experienced a treatment effect of 11%, significantly above the average treatment effect (ATE) of 1.6%. The red bars represent features that contribute positively to the treatment effect, including a high National Institutes of Health Stroke Score (NIHSS), a neurological exam for stroke evaluation, Total Anterior Circulation Infarct (TACI), and usage of anti-platelet medications within 48 hours; the blue bars indicate features that reduce the treatment effect, including atrial fibrillation history and higher systolic blood pressure. This demonstrates how LIFT-XAI decomposes the individual treatment effect into feature-level contributions, enabling detailed interpretation of clinical features for each patient. Similarly, the lower force plot shows a male patient with low NIHSS scores and Partial Anterior Circulation Infarct (PACI) syndrome, who experienced a negative treatment effect.

Figure 3 (c) illustrates individual feature attributions and each feature’s global ranking across cohorts, showing that NIHSS is the most influential factor affecting rt-PA efficacy. Without categorizations or creating numerous subgroups, we can easily examine the impact of continuous features. For example, Shapley value indicates that patients with higher NIHSS, depicted by the red cluster, contribute to treatment effects positively when administered TPA, in contrast to those with lower NIHSS scores, marked by the blue cluster. This observation is consistent with prior research ^24,30^, which also identified a significant interaction between NIHSS scores and rt-PA treatment effectiveness. Additionally, the second most impactful feature is the type or syndrome of the stroke. As shown in Figure 3 (c), rt-PA exhibits enhanced benefits for patients diagnosed with TACI and PACI, a finding consistent with the original IST-3 study and reported in several stroke-related studies ^31^. Our findings also reveal that factors such as receiving an anti-platelet drug within 48 hours and infarction history significantly affect the effect of rt-PA, which previous studies have also discovered ^31,32^.

With LIFT-XAI, we can perform subgroup analysis by grouping patients based on feature values and examining their Shapley contributions. This also enables us to explore feature interactions—how one variable affects another’s influence on treatment effect. In Figure 3 (d-top), we analyze the interaction between gender and NIHSS score. While NIHSS is a primary driver of rt-PA effect, LIFT-XAI reveals a gender-specific pattern: in females (red), high NIHSS scores contribute more positively, whereas low scores contribute more negatively. In contrast, the effect is less pronounced in males. A similar pattern is observed among patients without atrial fibrillation, where rt-PA appears significantly more effective in those with higher NIHSS scores, Figure 3 (d-bot).

### Use cases 2: clustering analysis on LIFT-XAI results reveals novel subgroup

We further explored explanation-based patient clustering in a subset of individuals from the IST-3 trial. Specifically, we focused on patients with NIHSS scores between 6 and 14—a group with uncertain treatment effects for or against rTPA treatment according to trial subgroup analysis, and no clear clinical guidelines. Conducting a more fine-grained analysis within this intermediate group may help reveal subtle differences among patient subpopulations and could yield new insights for clinical decision-making. To this end, we applied LIFT-XAI to examine this subgroup through their Shapley values and performed hierarchical clustering directly in the explanation space.

This approach provides deeper insights and highlights substantial heterogeneity in feature contributions, even for dominant variables like the NIHSS score. This indicates that similar clinical profiles based on a single feature do not necessarily imply similar treatment effect mechanisms, Figure 3 (e). Feature clustering reveals distinct patient clusters where features such as diastolic blood pressure and body weight contribute differently to the rT-PA effect, thus offering unique patient groups and potentially modifiable features that can be optimized to improve treatment effect where previous RCT’s suggest none exist, Figure 3 (f). (For details of feature distribution for each cluster, please refer to Table S.10.) By moving beyond feature-based subgrouping to explanation-based subgrouping, LIFT-XAI addresses the limitations of traditional subgroup analysis and uncovers deeper patterns of treatment effect heterogeneity.

### Use cases 3: deciphering treatment effects across RCTs when patients are different

A common reason why RCTs cannot be applied to more general populations is due to variation in patient characteristics that influence treatment effects. To address this issue, We stress-tested LIFT-XAI’s ability to identify key differences in patient characteristics driving alternative treatment outcomes in the setting of intensive blood pressure management using two notable RCTs: the SPRINT trial showed that intensive blood pressure management reduced cardiovascular events and mortality in high-risk, non-diabetic patients, whereas the ACCORD trial found no significant benefit when the same treatment was applied to patients with type 2 diabetes ^21,22^.

We first compared the top features affecting treatment outcomes in both trials. Interestingly, despite overall similarities between the cohorts, the top features affecting the treatment effect for each trial were quite different. In ACCORD, the most significant feature affecting the treatment effect was a *history of CVD*, followed by gender, aspirin use, number of antihypertensive medications, and an individual’s ethnicity, Figure 4 (a-top). Conversely, in the SPRINT trial, *age* was the most significant factor influencing blood pressure control, followed by gender, statin usage, chronic kidney disease history (CKD), and cardiovascular (CVD) history, Figure 4 (a-bot)

**Fig. 4.**
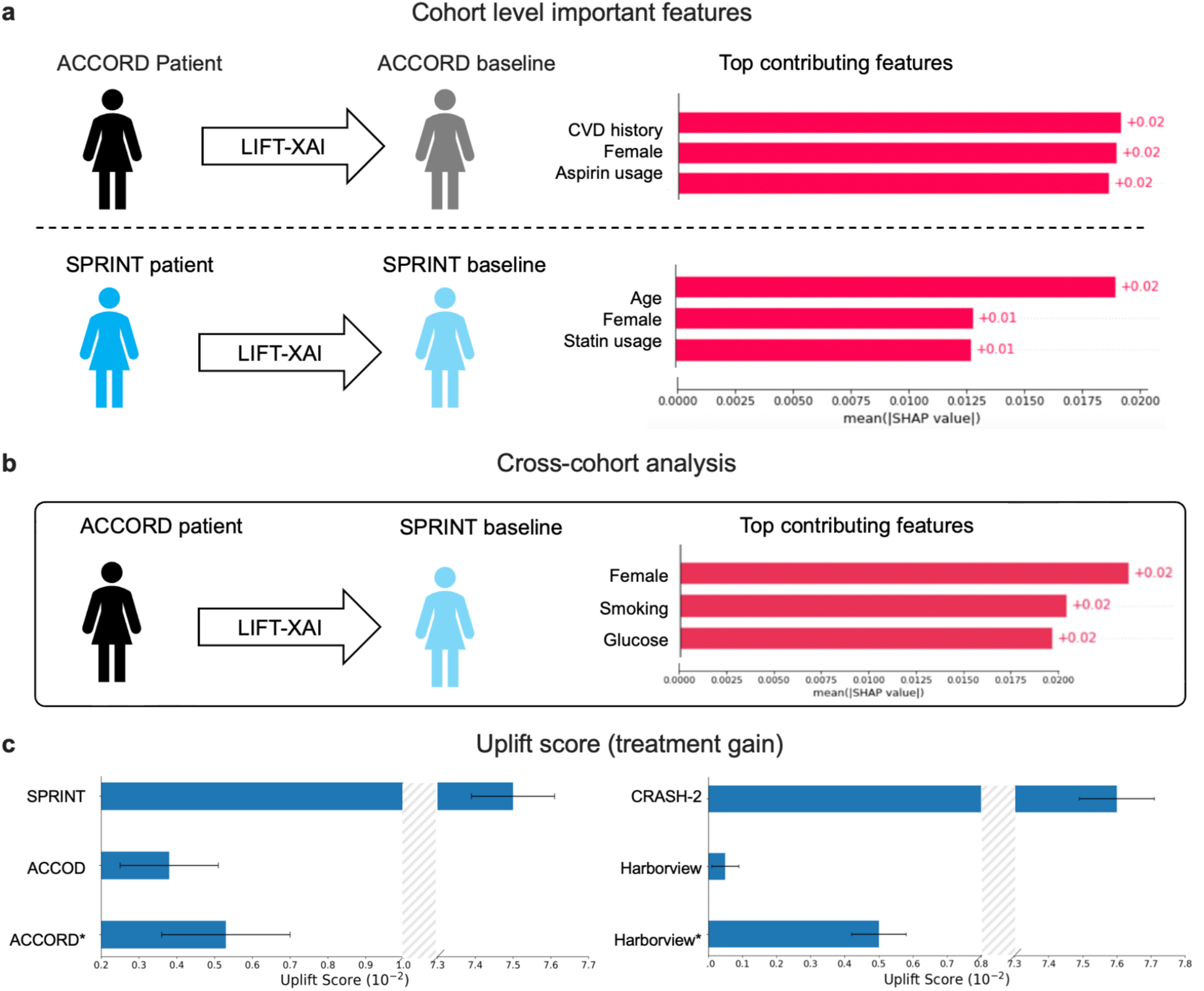
(a) Top 3 features based on LIFT-XAI’s global explanation in the ACCORD and SPRINT studies. (b) Analysing ACCORD patients with SPRINT baseline and its top contributing features through LIFT-XAI. (c) Uplift score (treatment gain) for SPRINT and ACCORD (left); CRASH-2 and Harborview trauma registry(right). (*) denotes patient population excluding individuals with glucose levels greater than 300 mg/dL in ACCORD and patients aged over 45 years in the Harborview trauma registry.

To enable further cross-cohort comparisons of ACCORD vs. SPRINT, we substituted the ACCORD baseline of diabetic individuals with representative non-diabetic individuals from the SPRINT cohort, Figure 4 (b); Section C.2. This reframes the original question—*which features are important for ACCORD individuals compared to the average ACCORD population?* —to instead ask *which features are important for ACCORD individuals compared to the average SPRINT population?* By adjusting the reference group accordingly, this approach supports tailored, context-specific interpretations across studies.

Upon reassessing the top features from both cohorts and reanalyzing the feature rankings, we observed that *fasting glucose (fpg) emerged as a prominent feature for diabetic patients in ACCORD, but it ranked 14th among the 18 clinical features for non-diabetic patients in SPRINT* ; see Figure S.8 (a). By identifying fasting glucose as a key treatment effect, LIFT-XAI correctly and independently identified the underlying key patient characteristic, i.e. the presence of diabetes, most likely driving the difference in treatment effect between the two trials. Moreover, LIFT-XAI independently provided a clear and usable treatment metric (fasting glucose) for clinicians seeking to manage blood pressure in diabetic patients, suggesting that intensive blood pressure management may actually be useful in diabetic patients if blood glucose is tightly controlled

To further investigate the impact of glucose on the effectiveness of blood pressure control in the ACCORD study, we analyzed the treatment uplift using qini scores and uplift scores (Section B.3) among patients with varying glucose levels. As we show in Figure 4 (c-left) and Table S.11, the uplift score and qini score for the original ACCORD was 3.8 *×* 10^−3^ and 2.2 *×* 10^−3^, respectively, significantly lower than the SPRINT studies, i.e., 7.5 *×* 10^−2^ and 3.9 *×* 10^−2^, respectively. However, when excluding patients with glucose levels exceeding 300 mg/dL (the maximum observed value in the SPRINT cohort), the average treatment effect of ACCORD increased by 39.5% for the uplift score and 36.3% for the qini score.

Using LIFT-XAI, we thus unravel these conflicting results in trials. Our analysis highlights variances in glucose levels as a potential explanatory and modifiable risk factor for the observed disparities in treatment outcomes between the two studies.

### Use cases 4: applying LIFT-XAI across clinical practice settings

Here, we test the ability of LIFT-XAI to identify important features contributing to treatment effects when a proven treatment is applied to a different clinical setting. For this test, we considered the treatment of traumatic bleeding after injury using tranexamic acid (TXA), a drug that is used to stabilize blood clots to reduce bleeding after injury. Strong randomized data favor the use of TXA for trauma victims at risk of significant bleeding if given at hospital admission and within 3 hours of injury ^25^. Time from injury has emerged as having an important effect on TXA efficacy. Consequently, clinical practice has steadily crept towards using this drug faster, either at the scene of injury or during transport (pre-hospital), despite the lack of randomized evidence for its efficacy in this alternative practice setting.

In this scenario, we asked LIFT-XAI to identify *which features were most important for trauma patients when TXA was given in the hospital setting vs. when TXA was given pre-hospital*. Using data made available from CRASH-2 study investigators ^25^ and our local trauma center registry, we asked LIFT-XAI to identify features that determine

TXA efficacy when administered in these different clinical practice settings (Section A.2). We then validated the feature selected by LIFT-XAI in the new pre-hospital setting by computing the treatment effect gain. To validate our results, we then compared them to features identified during a more recent randomized controlled trial of TXA when given specifically in the pre-hospital setting ^33^.

We first compared the top features based on LIFT-XAI. As shown in Figure S.10 (a-left), in the pre-hospital settings, the top features were time-to-injury, Glasgow Coma Score (GCS), trauma type, and a new effect, *age* that was not previously identified in CRASH-2 subgroup analysis. We then examined the treatment effects among different age groups. As shown in Figure 4 (c-right) and Table S.11, the uplift score and qini score for our pre-hospital cohort are 5 *×* 10^−4^ and −5 *×* 10^−4^, respectively. Surprisingly, after excluding patients aged over 45 years in the pre-hospital settings, the scores increase to 5 *×* 10^−3^ and 8 *×* 10^−4^, respectively. This finding indicates that, in the pre-hospital setting, LIFT-XAI is identifying age as a new and potentially crucial correlate of TXA efficacy. This result was validated by similar emergence of age as a new treatment effect for TXA efficacy from the PATCH study, a randomized controlled trial of TXA administered to injured patients in the pre-hospital clinical setting ^33^. This result highlights the ability of LIFT-XAI to identify important treatment effects when randomized clinical trial data are applied towards different clinical practice settings.

## Discussion

We demonstrate that providing a deeper understanding of CATE dynamics with XAI can extend the capability of RCTs to unveil real-world clinical insights and support physicians to make better-informed treatment decisions. Specifically, we present a framework, LIFT-XAI, that rigorously explains CATE ensemble models with Shapley values, improving our ability to realize how randomized controlled trial data can be applied to real-world treatment decisions in a robust manner. We demonstrated that ensemble CATE models can effectively estimate treatment effects using real-world clinical data in a controlled environment with proper model selection criteria. Furthermore, Shapley values consistently outperform alternative attribution methods in terms of local fidelity, global sufficiency, and explanation stability, making them particularly well-suited for interpreting the CATE estimates. When combined, these techniques strengthen the utility of feature attribution methods and CATE models, facilitating the integration of machine learning and XAI into clinical translation ^11^ and enabling a deeper understanding of the potential risks and benefits of new treatments for the individual patient.

LIFT-XAI enabled identification of key clinical features that could be used to modify treatment decisions across a range of both acute and chronic health conditions. ^21,22,24,25^ At the cohort level, LIFT-XAI was capable of independently explaining conflicting results between trials and identified key covariates. For example, the effect of intensive blood pressure management on health outcomes was studied in two RCTs, SPRINT and ACCORD. The trial results did not agree. The SPRINT RCT demonstrated benefit ^22^, however, the ACCORD RCT showed no benefit for a patient cohort that included those with diabetes mellitus ^21^, suggesting the presence of diabetes as having a confounding effect upon blood pressure treatment efficacy. We used LIFT-XAI to explain the treatment effects for these RCTs and it independently identified blood glucose concentration as having a primary influence on blood pressure treatment efficacy. This result is important because it both supports the evidence that intensive blood pressure treatment does not benefit those patients with diabetes mellitus, thus improving patient selection for this therapy. In addition, this result suggests that blood glucose management is a potential therapeutic maneuver that may be used to allow patients with diabetes to derive benefit from intensive blood pressure treatment once their blood glucose concentration is controlled. Importantly, we found that LIFT-XAI treatment effect explanations could be extended to individual patients, highlighting its potential to inform personalized treatment decisions.(Figure 3).

However, LIFT-XAI is constrained by inherent limitations in both CATE models and Shapley values. Although CATE estimators perform well in controlled settings such as randomized controlled trials (RCTs), their reliability is reduced in observational studies where unobserved confounders may bias effect estimates. Such confounding can violate key identification assumptions, compromising model validity ^6,11^ and propagating bias into the resulting explanations ^19^. Beyond model limitations, while feature attribution methods are valuable for highlighting candidate features of treatment effect heterogeneity, they cannot establish causal relationships ^34^ and are sensitive to input and model perturbation ^35^. Misinterpreting attribution scores as causal drivers may therefore lead to misleading inferences about treatment effect mechanisms. We therefore recommend using LIFT-XAI primarily as an exploratory tool for feature discovery, while emphasizing that rigorous clinical trials remain essential to validate clinically actionable insights.

A promising future research direction involves developing methods to impute robust attribution scores to mitigate the impact of these confounders ^36^. Additionally, integrating causal knowledge with domain expertise has been demonstrated to enhance the accuracy of feature attributions ^37^.

To conclude, we present a new approach to explaining treatment effects using LIFT-XAI. We propose evaluation methods to assess CATE models with XAI using the gold standard for determining treatment efficacy in medicine, randomized clinical trials. LIFT-XAI demonstrates several advantages compared to traditional subgroup analysis, including individual explanation, subpopulation analysis, and cross-cohort examinations. In an era where precision medicine and individualized treatments are taking center stage, understanding the nuances of treatment effects is more crucial than ever.

## Methods

### Datasets

Most prior works of interpretability methods for conditional average treatment effects (CATEs) have focused on semi-synthetic or non-clinical benchmarks, such as IHDP and ACIC ^14,19^. Only a few studies have examined clinical data, typically restricted to a single trial with modest sample sizes ^12,20,38^, leaving evaluation in real-world clinical settings underdeveloped.

To address this gap, we analyzed four publicly available randomized controlled trials (RCTs) with binary outcomes. These trials were chosen for their clinical significance and their large sample sizes, making them a reliable testbed for CATE models. Randomization in RCTs not only reduces confounding but also provides the conditions needed for valid estimation of treatment effects ^2^. Each dataset was curated by removing variables with substantial missingness and applying one-hot encoding to categorical variables. The resulting datasets are:

#### IST-3

This trial evaluated intravenous alteplase (rt-PA) versus control in patients with acute ischemic stroke (*n* = 3,035)^24^. The dataset includes 7 continuous, 4 binary, and 1 categorical covariate (stroke type), with 6-month survival as the binary outcome.

#### CRASH-2

This trial tested tranexamic acid (TXA) against placebo in bleeding trauma patients (*n* = 20,211)^25^. The dataset includes 7 continuous, 1 binary, and 1 categorical (injury type) variables, with in-hospital survival defined as the binary outcome.

#### SPRINT

SPRINT ^22^(*n* = 9,361) compared intensive blood pressure control with a standard target . The dataset contains 12 continuous and 7 binary covariates, with the primary binary outcome defined as freedom from composite cardiovascular events.

#### ACCORD

ACCORD ^21^ evaluated intensive versus standard systolic blood pressure targets in patients with type 2 diabetes (*n* = 10,251). It includes 18 continuous and 7 binary covariates, with a protocol-defined primary cardiovascular composite outcome.

In particular, SPRINT and ACCORD enrolled overlapping patient populations but reported contrasting outcomes for intensive blood pressure control, offering a natural case study for whether interpretability methods capture clinically meaningful differences.

To complement these RCTs, we incorporated a retrospective cohort to assess performance in a real-world clinical setting. This retrospective cohort comes from the Harborview pre-hospital TXA registry (2007-2020) Section A.2, serving as a reference population for CRASH-2. We applied propensity-score matching ^39^ to construct a balanced sample of 240 patients (120 TXA, 120 controls) from trauma admissions. It contains 6 continuous and 1 categorical variable, with survival as the outcome. Detailed variable definitions of the above datasets are provided in Table 3.

**Table 3.** Curated dataset descriptions, including features, treatment, and outcome definitions.

| Dataset | Type (count) | Variables | Treatment | Outcome |
| --- | --- | --- | --- | --- |
| IST-3 | Continuous (7) | Age, Weight, Glucose, Glasgow Coma Scale (GCS) score<br>NIH Stroke Scale (NIHSS), Systolic Blood Pressure (SBP), Diastolic Blood Pressure (DBP) | Placebo<br>IV alteplase (rt-PA) | Alive and independent<br>at 6 months |
|  | Binary (4) | Sex, Antiplatelet use within 48 hours, history of Atrial fibrillation, Infarct present |  |  |
|  | Categorical (1) | Stroke type |  |  |
| CRASH-2 | Continuous (7) | Age, Systolic Blood Pressure (SBP), Respiratory rate, Capillary refill time<br>Heart rate, Time from injury to treatment, Glasgow Coma Scale (GCS) score | Placebo<br>Tranexamic acid (TXA) | In-hospital death<br>within 4 weeks |
|  | Binary (1) | Sex |  |  |
|  | Categorical (1) | Injury type |  |  |
| ACCORD | Continuous (12) | Age, Body Mass Index (BMI), Systolic Blood Pressure (SBP), Diastolic Blood Pressure (DBP)<br>Heart rate (HR), Fasting Plasma Glucose (FPG), Serum Creatinine (SCr), Estimated GFR (eGFR)<br>Urine Albumin-to-Creatinine Ratio (UACR), Alanine Aminotransferase (ALT),<br>Triglycerides (TG), Total Cholesterol (TC), LDL, HDL, Blood Pressure Medication Use | SBP ≤ 140 mm Hg (standard)<br>SBP ≤ 120 mm Hg (intensive) | Major CVD events |
|  | Binary (6) | Female, CVD history, Statin use, Aspirin use, Anticoagulant use, Current smoker |  |  |
|  | Categorical (1) | Race |  |  |
| SPRINT | Continuous (12) | Age, Systolic Blood Pressure (SBP), Diastolic Blood Pressure (DBP),<br>Estimated Glomerular Filtration Rate (eGFR), Serum creatinine (SCr), Total cholesterol (TC)<br>High-density lipoprotein (HDL), Triglycerides (TG), Urine Albumin-to-Creatinine Ratio (UACR)<br>Body Mass Index (BMI), Number of antihypertensive agents, Glucose | SBP ≤ 140 mm Hg (standard)<br>SBP ≤ 120 mm Hg (intensive) | Major CVD events |
|  | Binary (7) | Female, race, Current smoker, Aspirin use<br>Statin use, Subclinical cardiovascular disease (CVD), Chronic kidney disease (CKD) |  |  |
|  | Categorical (0) | Race |  |  |

### CATE Models

Under the potential–outcomes framework ^6^, the CATE for a binary treatment *T* ∈ {0, 1} and covariates *X* is

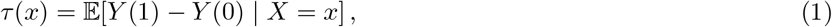

where *Y*_*i*_(1) and *Y*_*i*_(0) denote individual *i*’s potential outcomes under treatment and control. Because only one potential outcome is observed for each individual, *τ* (*x*) must be inferred from data. Several works have proposed neural network-based *meta-learners* ^2,19^ that estimate *τ* (·) by regressing learner-specific pseudo-outcomes:

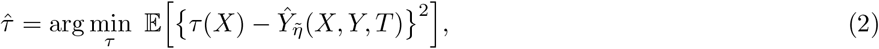

where 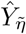 is constructed from nuisance functions 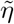. The specific form of 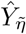 depends on the learner. We considered a broad family of learners, including S-, T-, X-, R-, RA-, and DR-learners, as well as TARNet, DragonNet, and CFRNet, and self-interpretable approaches, such as Causal Forests ^40^ and linear DR-Learners ^10^ (Section B.1). We implemented CATE estimators using the `CATENets` package^1^

#### Model training

Data were partitioned into training (70%), validation (20%), and test (10%) sets, with stratification by treatment (and outcome when binary) to preserve balance. Within the training set, we further applied stratified 5-fold cross-validation to obtain out-of-fold (OOF) predictions for model selection. Models were trained with early stopping, monitoring validation loss with a fixed patience parameter. Hyperparameters (learning rate, batch size, number of layers, hidden units, etc.) were tuned via grid search (Section B.2). For ensemble models, we trained 40 independent replicates with different random seeds and averaged predictions.

#### Evaluation

Because the true CATE *τ*^*^(*x*) is unknown, we evaluate models using *pseudo–outcome surrogates* ^41,42^ (Section B.3) that approximate the prediction error, as well as Qini and uplift curves ^43^ (Section B.3), which measure treatment effect gains based on the model’s estimates. Model selection uses 5-fold cross-validation, and the best-performing learner type is then designated as the *explainee* for subsequent interpretability analyses.

### LIFT-XAI: Explaining Ensemble CATE with Shapley Values

Reliable interpretability of CATE models is essential in clinical contexts, where explanations can yield patient-level insights in clinical decision-making. However, explanations are often based on a single model, which may not adequately approximate the underlying treatment effect ^44^. Prior works have also found that explanations from a single model are often unstable due to stochastic training factors such as random initialization and data shuffling ^18^, limiting their reliability as indicators of the true signal.

To address this, we propose explaining *ensemble models* ^45^. Concretely, we train *N* instances 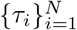 with distinct random seeds and data shuffling. Predictions and attributions are then averaged across models:

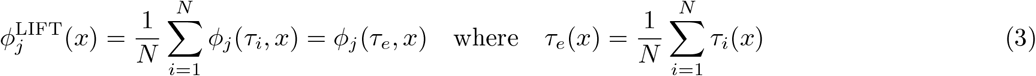

This equality holds for attribution methods that satisfy the *linearity* axiom (e.g., Shapley values, IG) (Section C.1). By reducing variance from stochastic training, this framework yields feature attributions that more robustly capture true treatment-effect signals.

#### Choice of attribution method

While multiple attribution methods could be employed to ensemble models, we adopt Shapley values ^15,16^ because they uniquely satisfy the axioms of *efficiency, symmetry*, and *linearity*, providing a theoretically grounded attribution. Prior studies have also shown their promise in identifying relevant features for treatment effect in semi-synthetic environments ^19,20^. Moreover, Shapley values extend naturally from individual-level to cohort-level explanations, aligning with both personalized and population-level clinical analyses. Formally, for features *F* and a model *τ* (*x*), the Shapley value of feature *j* for individual *x* is:

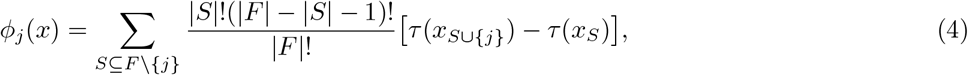

where *S* is a subset of features after excluding *j*. In practice, we can approximate Shapley values via sampling, using the empirical feature means as baselines to estimate marginal expectations^46^. This approximation directly addresses the clinically relevant question: *Which patient features most strongly drive deviations in treatment effect from what is expected for an average patient?* We therefore introduce **LIFT-XAI** as a principled framework for explaining ensemble CATE models using Shapley values, which we employ throughout this work.

### Examining Interpretability Methods

Because ground-truth feature contributions to treatment effects are rarely observable, we evaluate explanation methods using indirect but complementary criteria: (i) *local fidelity*, which tests whether attributions reflect model behavior under perturbations; (ii) *global sufficiency*, which examines whether the most important features identified by the explanation are sufficient to recover model predictions; and (iii) *consistency*, which measures the stability of explanations across independently trained models. When external ground-truth signals are available, we additionally evaluate (iv) *feature rank concordance*.

#### Local fidelity

Local fidelity ^47^ measures how well feature attributions capture the model’s prediction behavior at the individual level. We adopt standard insertion-deletion benchmarks ^26^ with the pseudo–outcome surrogate (Section B.3) as the fidelity metric. This assesses whether removing or inserting features with high attribution values leads to the expected changes in predicted treatment effects.

#### Global sufficiency

Global sufficiency assesses whether features identified by an explanation capture the model’s predictive signal ^48^. We rank features by their attribution values,^2^ then train a student model 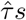 using only the top-*k* features to reproduce the ensemble’s predictions 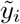.^3^ The distillation error over *M* test points is

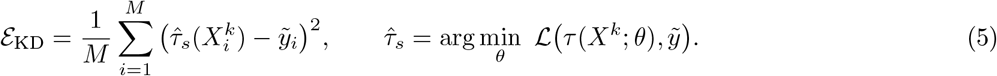

Lower ℰ_KD_ indicates that the explanation-selected features capture the teacher’s signal with fewer inputs.

#### Consistency

For explanations to be useful in scientific discovery, explanations must be stable across repeated training runs. We therefore assess consistency across *L* independently trained ensembles, 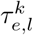 (with *L* = 1 corresponding to a single model), each comprising *k* base learners. We compute the mean pairwise cosine similarity of their *global* attribution vectors (average absolute attribution across individuals):

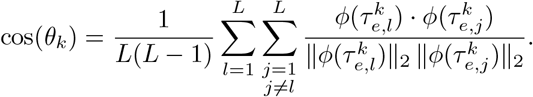

An increase in cos(*θ*_*k*_) with *k* indicates reduced sensitivity to random initialization and greater reflection of underlying signal rather than noise.

#### Feature rank concordance

When external ground-truth signals are available, we also assess rank concordance. Specifically, we compute *Spearman’s rank correlation* ^27^ between the global feature ranking from an explanation method, 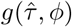, and oracle rankings *g*(*p*) derived from interaction *p*-values ^28^, where lower *p*-values denote stronger evidence of treatment effect heterogeneity. The resulting correlation

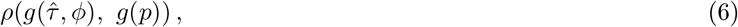

indicates whether explanation methods recover features consistent with ground truth features.

### Baseline Explanation Methods

We benchmark LIFT-XAI against widely used explanation methods at both the individual (local) and cohort (global) levels. At the individual level, we compare Shapley values to established attribution techniques, including Integrated Gradients (IG) ^49^, gradient-based Saliency maps ^50^, SmoothGrad ^51^, and LIME ^52^, which provide feature-level attributions for each patient. At the cohort level, we additionally benchmark treatment-effect–specific methods such as Leave-One-Covariate-Out (LOCO) ^12^ and PermuCATE^13^, which yield global importance scores but do not produce individual-level explanations. Implementation details and hyperparameter choices are provided in Section C.2.

## Data availability

The generation process for synthetic datasets is available on GitHub at https://github.com/AliciaCurth/CATENets. The IST-3 dataset is publicly accessible at https://datashare.ed.ac.uk/handle/10283/1931. The CRASH-2 dataset can be accessed at https://freebird.lshtm.ac.uk/index.php/available-trials/, with treatment allocations available upon request. Both the ACCORD and SPRINT datasets are available upon request at https://biolincc.nhlbi.nih.gov/home/.

## Code availability

The code for training, inference, and evaluation of the CATE models and XAI methods used in this study is available at https://github.com/suinleelab/LIFT-XAI. The code is distributed under the BSD 3-Clause License. The model weights are provided and intended for non-commercial use only.

## Acknowledgement

We extend our gratitude to the Trial Collaborators at the London School of Hygiene & Tropical Medicine Clinical Trials Unit (University of London) for sharing CRASH-2 Trial data, and to the researchers in the Lee lab for their valuable discussions.

## Ethics declarations

## Competing interests

The authors declare no competing interests.

## Supplementary Information

### A Datasets

### A.1 Real-World RCTs

**IST-3 (The Third International Stroke Trial)**^***24***^ was a large, multicentre, randomized controlled trial designed to determine whether a broader range of stroke patients could benefit from intravenous recombinant tissue plasminogen activator (rt-PA) administered within six hours of acute ischemic stroke onset. The trial enrolled 3,035 patients across 156 hospitals in 12 countries, who were randomly assigned to receive either rt-PA or standard non-thrombolytic care. Functional outcomes were assessed at six months using the Oxford Handicap Scale (modified Rankin Scale). The results demonstrated that, in selected populations, the benefits of early thrombolytic therapy outweighed the associated risks.

The final curated dataset comprised 7 continuous variables (`age`, `weight`, `glucose`, `gcs-score-rand`, `nihss`, `sbprand`, `dbprand`), 4 binary variables (`gender`, `antiplat-rand`, `atrialfib-rand`, `infarct`), and 1 categorical variable (`stroketype`). The binary outcome variable represents six-month survival (`aliveind6`). For detailed information on inclusion criteria, participant demographics, and variable definitions, please refer to the original study ^24^ and its publicly available dataset at IST-3@DataShare.

**CRASH-2 (Clinical Randomisation of an Antifibrinolytic in Significant Haemorrhage)** ^***25***^ had the objective to evaluate the effect of early administration of tranexamic acid (TXA) on mortality, surgical intervention, and transfusion requirements in trauma patients with, or at risk of, significant bleeding. The randomized trial encompassed over 20,000 trauma patients. The intervention was TXA versus placebo. The major outcomes of interest were death in hospital within 4 weeks post-injury, need for blood transfusion, and surgical interventions. The major discovery was that early administration of TXA reduced all-cause mortality without increasing the risk of vascular occlusive events. The curated analytic dataset comprised 9 covariates in total: 7 continuous (`iage`, `isbp`, `irr`, `icc`, `ihr`, `ninjurytime`, `igcs`), 1 binary (`isex`), and 1 categorical (`iinjurytype`, one-hot encoded for analysis). The binary outcome variable represented survival at hospital discharge. For detailed information on inclusion criteria, participant demographics, variable definitions, feature distributions, and trial methodology, please refer to its original study^25^ and the publicly available dataset at freeBIRD.

**SPRINT (Systolic Blood Pressure Intervention Trial)** ^22^ was a large, multicenter randomized controlled trial designed to assess the effects of intensive versus standard blood pressure control on cardiovascular outcomes and overall mortality in adults at elevated cardiovascular risk. The trial enrolled 9,361 participants with a systolic blood pressure of 130 mmHg or higher and at least one additional cardiovascular risk factor. Participants were randomly assigned to an intensive treatment group targeting a systolic blood pressure below 120 mmHg or a standard treatment group targeting below 140 mmHg. The primary composite outcome comprised myocardial infarction, non–myocardial infarction acute coronary syndrome, stroke, heart failure, or death from cardiovascular causes. The study demonstrated that intensive blood pressure control significantly reduced the rates of the primary composite outcome and all-cause mortality compared with standard treatment.

The dataset comprises 19 baseline variables: 12 continuous, 7 binary, and no categorical variables. The final analytic dataset includes 12 continuous variables (`age`, `sbp`, `dbp`, `n-agents`, `egfr`, `screat`, `chr`, `glur`, `hdl`, `trr`, `umalcr`, and `bmi`) and 7 binary variables (`female`, `race-black`, `smoke-3cat`, `aspirin`, `statin`, `sub-cvd`, and `sub-ckd`). The treatment variable (`intensive`) indicates the randomized intervention arm, and the outcome variable (`event-primary`) represents the occurrence of the composite cardiovascular endpoint. For detailed information on inclusion criteria, participant demographics, variable definitions, feature distributions, and trial methodology, please refer to its original study ^22^.

**ACCORD (Action to Control Cardiovascular Risk in Diabetes)** ^21^ was a large, multicenter randomized controlled trial designed to evaluate the effects of intensive control of blood pressure, glucose, and lipids in adults with type 2 diabetes. The trial enrolled 10,251 participants who were randomly assigned to various intervention arms, including intensive versus standard blood pressure control (targeting below 120 mmHg versus below 140 mmHg, respectively). The primary composite outcome was the first occurrence of nonfatal myocardial infarction, nonfatal stroke, or death from cardiovascular causes. Results indicated that intensive blood pressure control did not significantly reduce the rate of the primary outcome compared to standard treatment. However, intensive glucose control reduced the rate of certain cardiovascular events while increasing overall mortality, and specific lipid management strategies demonstrated modest benefits ^21^.

The curated dataset from this trial comprises 18 continuous variables (`baseline-age`, `bmi`, `sbp`, `dbp`, `hr`, `fpg`, `alt`, `cpk`, `potassium`, `screat`, `gfr`, `uacr`, `chol`, `trig`, `vldl`, `ldl`, `hdl`, and `bp-med`), 7 binary variables (`female`, `cvd-hx-baseline`, `statin`, `aspirin`, `antiarrhythmic`, `anti-coag`, and `x4smoke`), and 1 categorical variable (`raceclass`).

These variables represent baseline demographic, biochemical, and treatment-related measures used to assess the effect of intensive interventions on cardiovascular outcomes. The treatment variable (`treatment`) indicates intensive versus standard blood pressure control, and the outcome variable (`censor-po`) represents the primary composite cardiovascular event. For detailed information on inclusion criteria, participant demographics, variable definitions, feature distributions, and trial methodology, please refer to its original study ^21^

For ACCORD and SPRINT, both datasets are available at BioLINCC repository upon request.

#### A.2 Observational Data

##### Harborview pre-hospital TXA cohort

The emergency medicine datasets used in this study were gathered over 13 years (2007-2020) and encompass 14,463 emergency department admissions. It is the only retrospective electronic health record (EHR) data that we used in our study. We excluded patients under the age of 18 and patients with hypotension, and we curated a clinical cohort with patients prescribed tranexamic acid (TXA) among trauma patients and the corresponding control group. The corresponding cohort group is selected based on propensity score matching ^39^. The cohort consists of 240 patients with 120 in the treated group and 120 in the control group. We selected variables similar to CRASH-2 settings, including trauma type, demographic information (age, sex), and pre-hospital vital signs (blood pressure, heart rate, respiratory rate). The outcome was each patient’s survival. This dataset is not publicly available due to patient privacy concerns.

#### A.3 Data Preprocessing

For our analysis, we curated each dataset by excluding records with substantial missingness (greater than 90%). For variables with missingness below this threshold, missing values were imputed using the empirical mean via the `SimpleImputer`function from the *scikit-learn* library. All continuous features were then normalized to the range [0, 1] using min–max scaling to ensure comparability across variables.

We also aggregated and removed redundant or overlapping features to simplify the feature space based on consultation with licensed physicians. For example, the Glasgow Coma Scale (GCS) for eye, motor, and verbal responses were combined into a single total GCS score. Additionally, highly correlated variables, such as the NIHSS score and its predicted counterpart, or overlapping indicators of stroke presence and symptom severity, were excluded.

#### A.4 Semi-synthetic Data

In our synthetic data environment, we replicate the data generation process described in ^19^. This approach generates semi-synthetic data based on the Twins, News, and ACIC2016 datasets. It’s important to note that our primary focus is on varying confounding settings. Within these settings, treatment assignment is determined by:

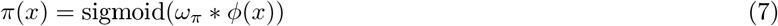

In this equation, *ω*_*π*_ stands for the confounding (propensity) scale, and *ϕ*(*x*) represents the confounding function. The confounding is either predictive, with *x* = *I*_*pred*_, prognostic, defined by *x* = *I*_*prog*_, or irrelevant, where *x* = *I*_*irrelevant*_.

##### Twins

The Twins dataset ^53^ encompasses 11,400 twin births in the USA from 1989 to 1991. It includes 39 covariates—both continuous and categorical, related to parents, pregnancy, and birth details.

##### News

The News dataset comprises 10,000 randomly sampled news items, each characterized by 2,858-word counts ^8,54^. The dataset is processed with Principal Component Analysis, with the first 100 principal components serving as continuous covariates for each item.

##### ACIC2016

The ACIC2016 dataset consists of data from the Collaborative Perinatal Project provided as part of the Atlantic Causal Inference Competition (ACIC2016) ^55^. It consists of 55 mixed (continuous and categorical) features for 2,200 patients.

### B Potential Outcome Framework

Using the Neyman–Rubin potential outcome framework ^6^, suppose a superpopulation 𝒫 gives rise to a sample of *N* independent random variables, represented as (*Y*_*i*_(0), *Y*_*i*_(1), *X*_*i*_, *W*_*i*_) ∼ 𝒫. Here: - *X*_*i*_ ∈ ℝ^*d*^ denotes a *d*-dimensional feature vector. - *W*_*i*_ ∈ {0, 1} signifies the treatment assignment, with the specific meaning to be clarified later. - *Y*_*i*_(0) and *Y*_*i*_(1) are the potential outcomes for unit *i* when assigned to the control and treatment groups, respectively. From this, we denote the Average Treatment Effect (ATE), as:

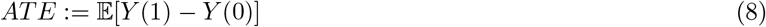

A core challenge in causal inference is the inability to observe both potential outcomes for each unit. For each unit, either the outcome under control (*W*_*i*_ = 0) or under treatment (*W*_*i*_ = 1) is observed, but not both. Given this, the observed data is:

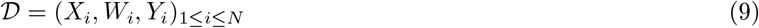

To decide the treatment for a new individual *i* with covariate *x*_*i*_, we aim to estimate its Individual Treatment Effect (ITE), *D*_*i*_ = − *Y*_*i*_(1) − *Y*_*i*_(0). However, since *D*_*i*_ remains unobserved and is not identifiable without robust assumptions, we focus on estimating the Conditional Average Treatment Effect (CATE), *τ* (*x*):

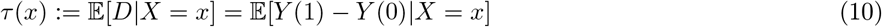

Note that the optimal estimator for CATE is also the best for ITE in terms of Mean Squared Error (MSE) ^23^. We aim for estimators that minimize the Expected Mean Squared Error (EMSE) for CATE estimation. This metric is also called the *precision in estimating heterogeneous effects (PEHE)*. ^56^.

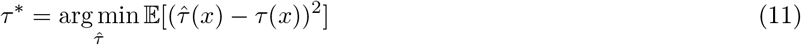

To be able to identify the causal effects from observational data, we make the standard assumptions for both domains. **Assumption 1**. *(Unconfoundedness)* There are no unobserved confounders, such that the treatment assignment and POs are conditionally independent given the covariates:

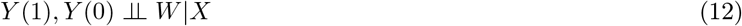

**Assumption 2**. *(Overlap)* for all *x* ∈ ℝ^*d*^,

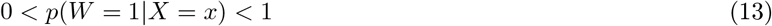

#### B.1 CATE Models

##### B.1.1 Meta Learners

In this section, we provide the background of meta-learners mentioned in the main text.

###### S-learner (Single Learner)

The S-learner trains a single model on both treated and control units.

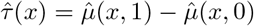

where 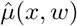 is the prediction of the model given covariate *x* and treatment *w*.

**T-learner (Two Learner)** ^23^ introduced the T-learner, an approach that creates distinct regression functions for each treatment group and computes the differences. The T-learner trains separate models for treated and control units.

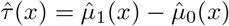

where 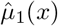 and 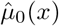 are the predictions for treated and control units, respectively.

**X-learner** ^23^ also introduce the X-learner, a two-step regression estimator that uses each data point twice. The X-learner is designed to leverage both the treated and control groups for estimating heterogeneous treatment effects. To start, models 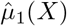 and 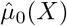 are fitted on the treated and control groups respectively. Using these models, treatment effects for the treated group are calculated as 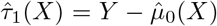 and for the control group as 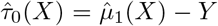. Subsequently, counterfactual outcomes for the treated group are estimated using *g*_0_ to get 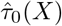 and for the control group using *g*_1_ to get 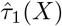. The final treatment effect estimate integrates these results using the propensity score, *e*(*X*) = *P* (*W* = 1|*X*)

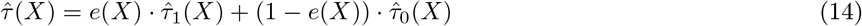

**DR-learner (Doubly Robust Learner)** ^10^ introduced the DR-learner, or doubly robust learner, a two-step procedure that employs the formula for the doubly robust augmented inverse propensity weighted (AIPW) estimator ^57^ as a pseudo-outcome in a two-step regression framework. The DR-learner combines propensity score weighting with regression adjustment.

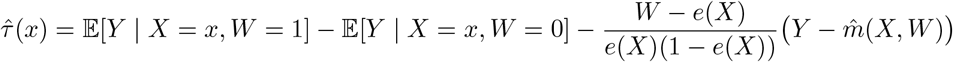

where *e*(*X*) is the propensity score.

**R-learner** ^58^ proposed the R-learner, which demands an estimate of the treatment-unconditional mean. The R-learner utilizes residuals to estimate the heterogeneous causal effect. Given a model, the residuals are computed as 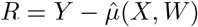. The causal effect is then estimated as:

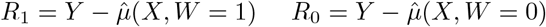

The causal effect is deduced from the relation between the residuals and the treatment assignment, given by:

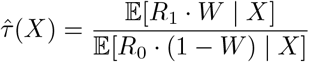

In this approach, the essence is to determine how deviations from the model’s predictions (residuals) correlate with the treatment, adjusted by the propensity of receiving the treatment.

##### B.1.2 Representation Learners

In addition to meta-learners, recent studies aim to learn a covariate shift function ^8^, ensuring that the feature representation of both treated and untreated groups have similar distributions. Algorithms such as CFRNet ^8^, TARNet ^59^, and Dragonnet ^60^ are introduced to predict CATE using balanced representations derived from observational data.

Specifically, the balanced learning approach identifies a representation, *h* : 𝒳 → ℛ, and treatment-specific functions, *µ*_1_ and *µ*_0_, which aim to minimize the PEHE evaluation measure. The model is trained using a loss function that’s bounded by PEHE. This function comprises the combined expected factual treated and control losses for outcome regression and the distance between *h*(*x*) for given *W* = 1 and *W* = 0 values concerning the covariate shift.

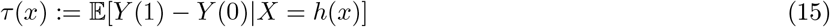

###### Dragonnet

Dragonnet employs a neural network architecture where both the treatment assignment and the outcome are jointly modeled. The network is structured to provide a representation *h*(*X*) of the input covariates that captures the nuances needed to predict both treatment propensity and potential outcomes. By jointly modeling, Dragonnet ensures that the learned representations are informative about both treatment assignments and outcomes.

###### TARNet (Treatment Agnostic Representation Network)

TARNet aims to learn a shared representation *h*(*X*) of the covariates for both potential outcomes. Once this representation is determined, it is then passed through two separate outcome models to predict the potential outcomes under treatment and control:

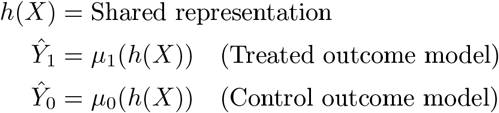

The shared representation ensures that the outcome predictions are based on a consistent understanding of the covariates.

###### CFR (Counterfactual Regression)

CFR focuses on deriving a shared representation *h*(*X*) that can produce balanced representations of treated and control units. This balance ensures that the distributions of the treated and control units in the representation space are similar, minimizing the distributional shift and aiding in counterfactual prediction. With this balanced representation, potential outcomes are then estimated using respective outcome models.

###### DR-CFR (Doubly Robust Counterfactual Regression)

DR-CFR combines the strengths of doubly robust estimation methods with the representation learning of CFR. After learning a balanced representation *h*(*X*), DRCFR not only predicts potential outcomes but also adjusts for discrepancies using propensity scores or outcome residuals, leading to a more robust and accurate estimate of treatment effects.

#### B.2 Model Implementation & Hyperparameters

For CATE model implementations, for S-Learner, T-Learner, X-learner, DR-learner, Dragonnet, TARNet, CFR, DRCFR, we rely on `CATENets`package^4^. For CausalForest and Linear DR-Learner, we follow the implementation from ^5^. Each nuisance function (i.e., 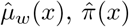, and 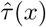) was constructed with the same number of hidden layers and hidden units across models. For architectures such as TARNet and CFRNet, the representation network Φ and the outcome heads *h*_*w*_ each contained one hidden layer with dense units and ReLU activations. For hyperparameter selection, we performed a grid search over the following ranges: training epochs, learning rates, batch sizes, numbers of hidden layers, and hidden dimensions. All models were trained using the Adam optimizer with early stopping based on validation performance.

#### B.3 CATE Model Evaluation

Here we introduce different evaluation strategies to validate CATE models.

**Factual criteria**. ^42^ evaluate models by measuring simple prediction loss only on the observed potential outcomes.

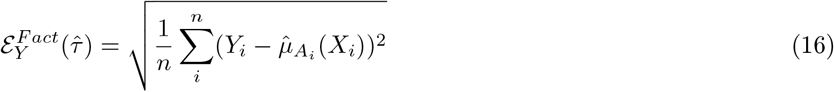

Obviously, one obvious disadvantage is that it may wrongly prioritize good fit on the potential outcomes over good CATE fit, resulting in bias ^42^.

##### Plug-in criteria

Construct surrogates for CATE evaluation by fitting a new CATE estimator 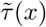 on held-out data and using this to compare against the estimates.

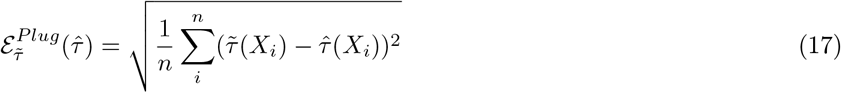

However, one obvious disadvantage for a plug-in surrogate is that it would favor models with similar structures e.g. S-Learner and T-Learner, a phenomenon called congenital bias. ^19,42^

##### Pseudo-outcome surrogate criteria

Obtain estimate through given auxiliary nuisance estimates 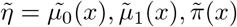 obtained from the validation data using ML method M, one can construct pseudo-outcomes 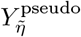 for which it holds that for ground truth nuisance parameter *η*, 𝔼 [*Y*_*η*_| *X* = *x*] = *τ* (*x*) and – instead of using them as regression outcomes as in the learners themselves

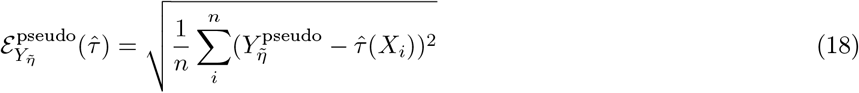

Here 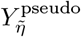 can be any pseudo-outcome objectives. In this work, we employ *Influence function, DR-Learner* objective, and *R-Learner* objective. Proposed by ^10^, DR pseudo-outcome employs doubly robust AIPW estimator and is hence unbiased if either propensity, 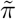, or outcome regressions, 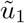 and 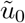, are correctly specified. 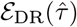 and 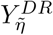 are denoted as,

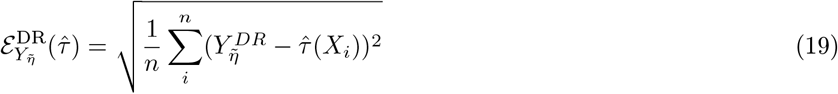

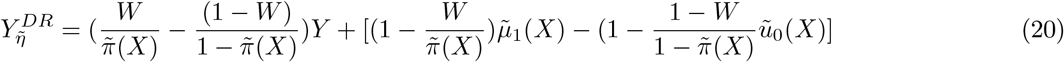

The R-learner objective of ^58^, which requires an estimate of the treatment-unconditional mean *µ*(*x*) = *E*[*Y*| *X* = *x*], relies on a similar idea and can also be used for the selection task, resulting in the criterion.

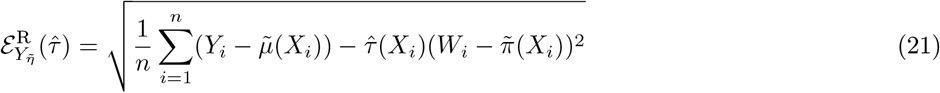

The influence function objective ^41^, categorized as surrogate pseudo-outcome ^42^, leverages Talyor-like expansion to approximate the PEHE.

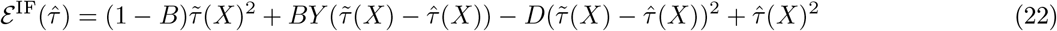

where 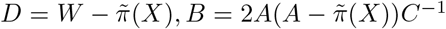 and 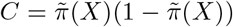, and 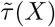 is a plug-in estimate. Surrogate pseudo-outcome is an unbiased estimator for CATE and more robust to congenital bias ^42^. On the theoretical side, the analysis in ^61^ shows that – under certain technical conditions – minimizing the surrogate validation estimate is a consistent model selection rule.

##### Qini Score and Uplift score

While pseudo-outcome surrogate provides an estimation for model selection, evaluating the performance of the chosen model on a different cohort can be difficult. As it requires training a second CATE model and nuisance functions. Therefore, the Qini curve is often used to evaluate model performance across different cohorts. It is defined as follows:

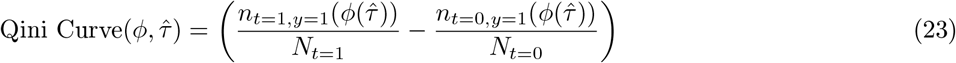

where 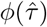 is the fraction of the population treated (in either treatment *t* = 1 or control *t* = 0 ) ordered by the uplift (treatment effect) from the model, 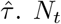 represent the total individual count in each group. Similarly, the uplift score can also be used to evaluate CATE model but only considering the treatment uplift within *ϕ* population. The uplift score is calculated as:

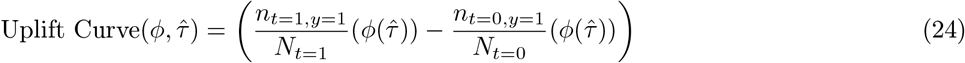

A baseline method is a model that cannot distinguish positive and negative uplift, and rank patients at random^6^. To quantify uplift/treatment effect from a given model, it is common to measure the area under the curve (AUROC) between the Qini/Uplift curve (intervention assignment based on model predictions) and random choice.

### C Interpretability (XAI) Methods

#### C.1 Key Explanation Properties for CATE Models

Considering the CATE setting, ^19^ suggests the following key properties for explanation methods, *a*_*i*_, *i* ∈ [*d*], in a CATE model.

##### Sensitivity

The covariates that do not affect the CATE model are given zero contribution. More formally, if for some *i* ∈ [*d*] we have 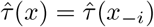 for all *x* ∈ *X*, then 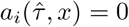 for all *x* ∈ *X* .

##### Completeness

Summing the importance scores gives the shift between the CATE and a baseline. More formally, for all *x* ∈ 𝒳, we have:

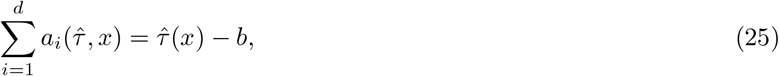

where *b* ∈ *R* is a constant baseline. In this way, each importance score *a*_*i*_ can be interpreted as the contribution from covariate *i* of*x* to have a CATE that differs from the baseline *b*. Note that the choice of the baseline differs from one method to another. For instance, the baseline for SHAP ^16^ is the average treatment effect: 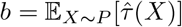.

##### Linearity

The importance score of a covariate is linear with respect to a black-box function. If the CATE model 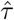 is written in terms of the estimated potential outcomes 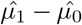, it can be written as 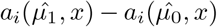. This makes it easy to differentiate prognostic and predictive covariates. If *x*_*i*_ is a prognostic covariate, then 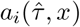 is zero. If *x*_*i*_ is a predictive covariate, then 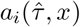 is not zero.

##### Model Agnosticism

The feature importance score can be computed for all CATE 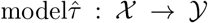. Some methods only work with a restricted family of models, which prevents them from being model agnostic. For example, Gradient-based explanation methods ^50,62^ only work for 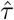 that is differentiable with respect to its input.

##### Implementation Invariance

The feature attribution would be the same for two functionally equivalent models. This means that if we have two CATE models 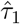 and 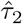 such that 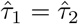 for all *x* ∈ 𝒳, this implies that 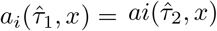 = for all *x* ∈ 𝒳 and all *i* ∈ [*d*].

#### C.2 Explanation Methods

In this study, we utilize explanation methods that fulfill the following criteria, including *sensitivity, completeness, linearity*, and *implementation invariance*. Accordingly, we employ Integrated Gradients ^49^ and Shapley values ^16^. Although Vanilla Gradient (Saliency) ^50^ doesn’t satisfy the completeness criterion, it’s served as a basic baseline. All the feature importance methods are implemented using Pytorch and Captum Python package^7^.

##### Vanilla Gradient

The vanilla gradient, often referred to as the saliency map ^50^, is frequently used as a benchmark for different explanation techniques. It is derived from the gradient of the output function in relation to the input features. Specifically, within CATE models, it assesses how the estimated treatment effect changes with respect to a given feature *x*_*i*_.

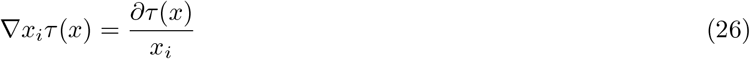

##### Integrated Gradients

Integrated Gradients (IG) assigns importance to input features by approximating the integral of a model’s gradients from a baseline input to the actual input ^49^. This method provides a holistic view of feature contributions, satisfying key properties like completeness. The IG attribution for an explicand *x*, a variable *x*_*i*_, and a baseline *x*^*′*^ is:

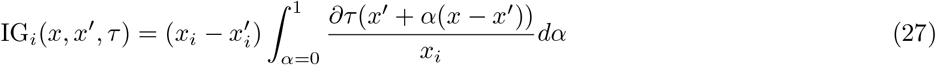

We use the zero vector as the baseline, denoted as *x*^*′*^ = 0. This means feature contributions are measured relative to their absence.

##### Shapley Value Sampling

Shapley Value, a concept borrowed from cooperative game theory, offers a unique approach to feature attributions ^16^. For any prediction model, it assigns each feature an importance value by averaging all possible combinations of feature presence or absence. Mathematically, for a prediction model *τ*, the exact Shapley value for a feature *x*_*i*_ is defined as:

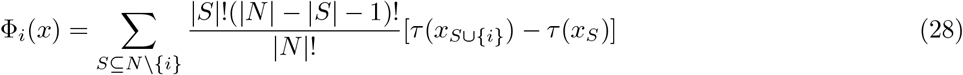

Where *N* is the set of all features and *S* is any subset of *N* that does not include feature *x*_*i*_. This equation evaluates the contribution of the feature *x*_*i*_ by contrasting the prediction with and without the feature over all possible combinations. However, computing the exact Shapley value can be computationally intensive, especially for models with a large number of features. Therefore, in practice, an approximation method like Shapley Value Sampling ^63^ is often used.

Given a feature set *N*, and for a particular feature *x*_*i*_, the sampled Shapley value is estimated as:

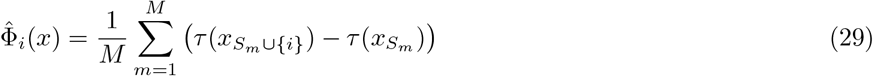

Where *M* is the number of sampled orderings and *S*_*m*_ is a random subset of *N* without feature *x*_*i*_ in the *m*^*th*^ sampled ordering. By sampling, we approximate the Shapley value, making it feasible for practical applications while maintaining a close approximation to the exact value.

##### Baseline Shapley

To obtain explanation/feature attribution for CATE models, we used the Shapley value ^15,16^, which is the average expected marginal contribution of adding one feature to the treatment effect after all possible combinations of features have been considered. More formally, the Shapley value takes as input a set function *v* : 2^*N*^ → *R*. The Shapley value produces attributions *s*_*i*_ for each player *i* ∈ *N* that add up to *v*(*N* ). The Shapley value of a player *i* is given by:

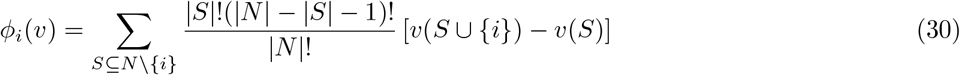

In other words, the Shapley value of player *i* is the average weighted average of its marginal contribution *v*(*S* ∪ {*i*} ) − *v*(*S*). Since computing the exact shapley value is intractable, several approximation methods have been proposed ^16,63^. Also, different choices of set function could lead to different Shapley values *ϕ*(*i*). One choice is to define *v*_*f,x*_(*S*) as the conditional expected model output on a data point when only the features in *S* are known ^46^

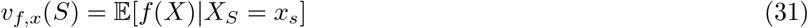

However, computing conditional distribution, 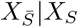, is usually difficult. Empirically, methods such as KernelSHAP ^16^ and shapley value sampling ^63^, conditional expectations are estimated by assuming feature independence; samples of the features in 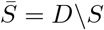 are drawn from the marginal joint distribution of these variables.

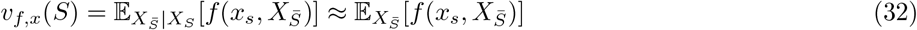

Moreover, we also leverage the assumption of model linearity to simplify the computation of the expected values ^16^. Empirically, we approximate the marginal distribution with an empirical mean of features as our baseline.

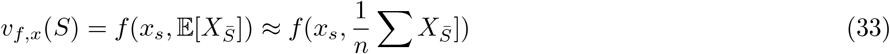

This approach is called Baseline Shapley (BShap) ^46^ and is more computationally efficient. More generally, this approach models a feature’s absence using its value in the baseline *x*^*′*^.

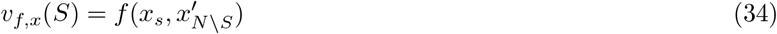

Another extension from baseline Shapley is Random Baseline Shapley (RBShap) ^46,64^ where the baseline 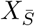 is drawn randomly according to the data distribution *D*. When propagating Shapley values, it approximates equation 32. However, we observed that this approach requires a significant amount of time to reach convergence. Therefore, in this work, we utilize BShap to explain CATE models.

##### Implementation details

For post-hoc interpretability at the individual level, we compared Shapley values with alternative attribution methods, including Integrated Gradients (IG) ^49^, gradient-based Saliency maps ^50^, Smooth-Grad ^51^, and LIME ^52^. All methods were implemented in `Captum`(v0.6.0) with respect to the CATE prediction 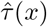. For Saliency, we used the `Saliency`class without smoothing; for IG, a mean baseline and 50 steps; for SmoothGrad, 50 noisy samples with Gaussian perturbations (*σ* = 0.2) averaged with `NoiseTunnel`; for Shapley, 500 permutations with the mean input as baseline; and for LIME, 500 perturbed samples with kernel width 0.75.

To obtain global feature rankings, we aggregated local attributions by taking the mean of their absolute values across individuals. For ensembles, attributions were computed for each model separately and then averaged across models.

#### C.3 Examination of Explanation Method in CATE

##### Predictive Features Identification with Ground Truth

Following the works of ^19,65^, within a semi-synthetic environment, the health outcome, denoted as *µ*_*W*_, can be expressed as a function of the patient characteristics *X* and the treatment *W* :

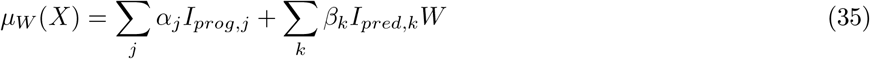

Where *I*_*prog,j*_ and *I*_*pred,k*_ denote the *j*^*th*^ and *k*^*th*^component of *X* that are prognostic and predictive covariates respectively. *α*_*j*_ and *β*_*k*_ are the corresponding coefficients. Under this assumption, the treatment effect can then be defined as:

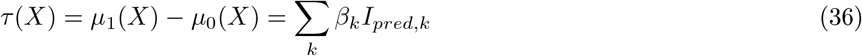

Therefore, in the context of CATE estimation, an explanation method should differentiate between predictive and prognostic covariates. To evaluate this, we can compute the average proportion of attribution correctly assigned to the predictive covariates:

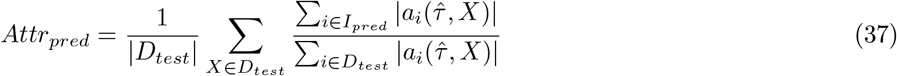

In this equation, *a*_*i*_ denotes the attribution score for a given feature *i*. This metric principally assesses the ability of an explanation method to differentiate between predictive and prognostic factors. While actual explanations or ground truths are absent in counterfactual predictions, this approach can offer guidance when choosing which explanation method is best suited for real-world datasets.

##### Ablation and Insertion and Deletion with Pseudo-outcomes

In practical scenarios, it’s usually impractical to obtain oracle features, therefore, to overcome this, one way is to perform an ablation test with CATE models. The ablation test is a commonly used evaluation technique for attribution values ^48^. Its concept involves replacing features with baseline feature values based on their attributions to measure their impact on the evaluation metric. This can be iteratively described using modified versions of the original explicands. For a given mixture of explicands, denoted by 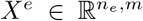, the attribution score is represented as *ϕ*(*f, X*^*e*^). The ablation study is characterized by three distinct parameters: (1) feature ordering, (2) an imputation sample, represented as *x*^*b*^ ∈ ℝ^*m*^, and (3) an evaluation metrics.

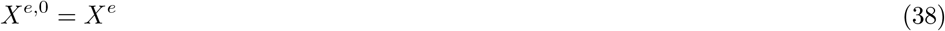

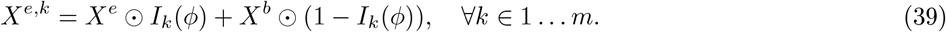

Where *X*^*b*^ := [*x*^*b*^ … *x*^*b*^]^*T*^ and *I*_*k*_(*ϕ*(*f, X*^*e*^)) = arg max_*k*,axis=1_(*ϕ*(*f, X*^*e*^)). The latter yields an indicator matrix the same size as *G*. A value of 1 indicates its position among the maximum *k* elements across a specific axis. The operation ⊙ represents the Hadamard product. Further, to assess the results of the ablation test, we compute the mean CATE output. When *k* features are ablated, this average is given by:

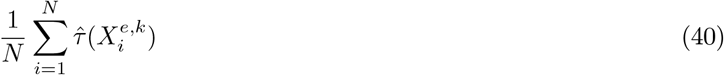

To contextualize the ablation test using treatment effects, imagine these features represent different features in a clinical trial, and their attributions (*ϕ* indicate the importance of each feature in predicting a particular effect of treatment, say patient recovery. When employing ablation: (1) Positive ablation: If we ablate (remove) features with the most positive attributions (those believed to have the most significant positive effect), we’d anticipate a substantial drop in, for example, patient recovery rates (mean model output). As we progressively remove additional features, this rate will continue to decline but at a diminishing pace. In this scenario, steeper declines imply that our attributions of features’ contributions are accurate. (2) Negative ablation: on the other hand, when removing features with the most negative attributions (those believed to hinder recovery), we’d expect recovery rates to increase significantly. More steep inclines would indicate better attributions, suggesting that the dropped features were indeed important.

In addition to CATE output, we can also leverage pseudo-outcome surrogates Section B.3, measuring the estimated PEHE when features are removed.

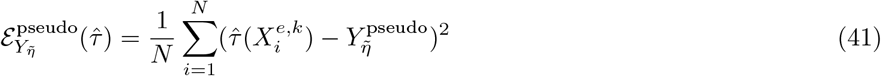

The term 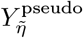 refers to the pseudo-outcome, such as the R or DR objectives, further detailed in Section B.3.

In addition to analyzing positive or negative ablations, insertion and deletion tests provide another way to assess performance. To conduct an insertion test, start with a baseline where no features are present and then gradually add features based on their ranking. Similarly, when assessing recovery rates, positive attributions should improve them while negative ones should have a negative impact.

### D Additional Experiment Results

#### D.1 CATE Evaluation in Clinical Datasets

In evaluating the Conditional Average Treatment Effect (CATE), we employed the pseudo-outcome surrogate approach, Section B.3. The experimental results are aggregated from 50 iterations of each model, each initialized with unique random seeds. A significant variance was observed with the influence function-based surrogate outcome metric, *ϵ*_if-pehe_. Owing to this instability, our model selection was principally driven by the more robust metrics, *ϵ*_*DR*_ and *ϵ*_*R*_, aligning with the findings of ^42^.

Our evaluations revealed minimal disparity in terms of the pseudo-outcome surrogate metrics *ϵ*_*DR*_ and *ϵ*_*R*_. Given that a majority of our datasets are sourced from randomized control trials—except the pre-hospital TXA dataset—this is anticipated. Especially in scenarios where the confounding (propensity) scale approaches 0, we observed a consistent identification of significant features across various explanatory methodologies, corroborated by the work of Crabbe et al. ^19^. We also observed that, for representation learners, the computational time for explanation is significantly longer than CATE models. Given these considerations, the X-Learner was predominantly selected as the model to be interpreted throughout our experiments.

#### D.2 Evaluating Ensemble CATEs’ efficacy

The second criterion involved comparing the ensemble-derived average treatment effect (ATE) estimates with the reported ATE values from each trial. In well-controlled randomized trials, where randomization minimizes confounding, the average of CATE estimates should closely align with the ATE in the absence of significant effect modifiers ^2^. For IST-3 and CRASH-2, the CATE models produced estimates closely aligned with the reported outcomes ^24,25^ (see Table S.3). However, in the blood pressure control trials, i.e., SPRINT and ACCORD, the CATE models provided higher ATE estimates than those reported: 1.6% for SPRINT and 1.2% for ACCORD, compared to 0.54% and 0.22%, respectively. Moreover, the analysis reveals that SPRINT, which demonstrated the efficacy of intensive blood pressure control, and the ACCORD study, which showed no significant treatment effects, align with the reported findings. These findings show that ensemble CATE models capture treatment effects at the cohort level, particularly in trials with substantial treatment effects.

#### D.3 Insertion & Deletion Procedure with Pseudo-outcomes - Real-world Data

In this section, we evaluate explanation methods using standard *insertion* and *deletion* experiments (see Appendix C). Relevant features are extracted with respect to the pseudo-outcomes, *ϵ*_pseudo_. We compute the Area Under the Curve (AUC) using a random baseline and normalize it by the number of features for each dataset. Our results show that the Shapley value method achieves the highest normalized AUC in the ACCORD, SPRINT, and CRASH-2 datasets, indicating superior performance. In contrast, for the IST-3 dataset, LIME performs best in insertion, while the Shapley value performs best in deletion. We also find that using *E*_*IF*_ tends to be relatively unstable, exhibiting high standard deviation, whereas *E*_*DR*_ and *E*_*R*_ are more consistent and stable across experiments.

#### D.4 Knowledge Distillation as a Benchmark Test in Clinical Datasets

#### D.5 Identifying Important Features to CATE in Semi-synthetic Environment

In this section, we evaluate explanation methods’ abilities in identifying important features in semi-synthetic environments A where we have access to oracle predictive features. These tasks include (1) identification of predictive features, (2) ablation studies with pseudo-outcomes, and (3) knowledge distillation with identified features.

##### Feature Identification Decrease as Confounding Effect Increases

Within the synthetic environment, we evaluated the performance of different explanation methods, Section C.3. Our results indicate that the Shapley value with the mean baseline consistently outperforms Integrated Gradients (IG) across various confounding effects in all datasets. However, as confounding effects intensify, the performance of all methods declines in tandem with the decreasing performance of the CATE model, Figure S.5 (a).

##### Insertion & Deletion Procedure with Pseudo-outcomes

We examine explanation methods with proposed evaluation task –ablation tests utilizing pseudo-outcomes. Notably, despite disparities between *ϵ*_pseudo_ and *ϵ*, their abilities in differentiating the performance of explanation methods are consistent. Results of ablation studies with *ϵ*_pseudo_ align with the findings from the one with true *ϵ*, as shown in Figure S.5 (b).

For the insertion procedure, where we start with a baseline and sequentially insert features based on their local rankings, both *ϵ* and *ϵ*_*pseudo*_ stabilize at approximately 8 features for IG, BShap - 0, and BShap - mean. In contrast, they settle at around 13 features for the vanilla gradient. On the other hand, in the deletion procedure, which is characterized by the stepwise removal of features starting from the most crucial, we find that IG, BShap - 0, and BShap - mean reach a steady state at around 6 features. In comparison, the saliency gradients exhibit stability a bit later, approximately at the 10-feature.

However, it’s worth noting that in semi-synthetic setups, the performance differential between IG and baseline Shapleys is almost negligible. Furthermore, similar to findings in S.5 (a), the performance shapley values with different baselines are indistinguishable in semi-synthetic contexts.

##### Knowledge Distillation as a Benchmark Test

In this section, we utilize model distillation as a means to assess various explanation methods in terms of global feature identification. Specifically, every student model adopts a 2-layer MLP architecture and the same hidden dimension. For each explanation method, we select the top features based on the average absolute attribution score. The results of this approach are presented in Table S.9, showing the performance of the student models on the test set across synthetic environments with different confounding effects.

To ensure the robustness of our results and to identify potential inconsistencies in the student model, we begin by training the student with a feature count, *n*, equivalent to the full set of predictive features utilized during data generation. The results, as depicted in Table S.9, indicate Mean Squared Errors (MSE) of 0.007, 0.028, and 0.003 for the Twins, ACIC, and News datasets respectively, across all the explanation methods evaluated.

However, when the number of input features is halved from the original set of predictive features, an expected decline in the student models’ predictive performance emerges. Notably, the saliency method lags behind its counterparts IG, BShap, and BShap across all datasets. These observations are aligned with our ablation studies presented in S.5. An intriguing finding is that, despite BShap’s superior performance in identifying predictive features, Fig S.5 (a), there’s a negligible difference among most explanation methods regarding global feature selection, except the vanilla gradient approach.

#### D.6 IST-3

##### D.6.1 Subpopulation Analysis

Here we demonstrate the results of shapley value by only considering a subgroup, male and female population specifically. For example, to understand important features only within the male population, for each feature, we used the average feature of males as the baseline for replacement during shapley calculation, Section C.2.

##### D.6.2 ACCORD & SPRINT Additional Results

Here, we present the feature rankings derived from Shapley values for the ACCORD and SPRINT cohorts. The Shapley values are calculated based on their respective population baselines.

##### D.6.3 Cross validation ACCORD with SPRINT

In this section, we demonstrate the CATE model’s performance in SPRINT (train) and ACCORD (test) with qini and uplift scores.

#### D.7 Pre-hospital (UW Harborview) & In-hospital (CRASH-2) TXA Additional Results

**Fig. S.1.**
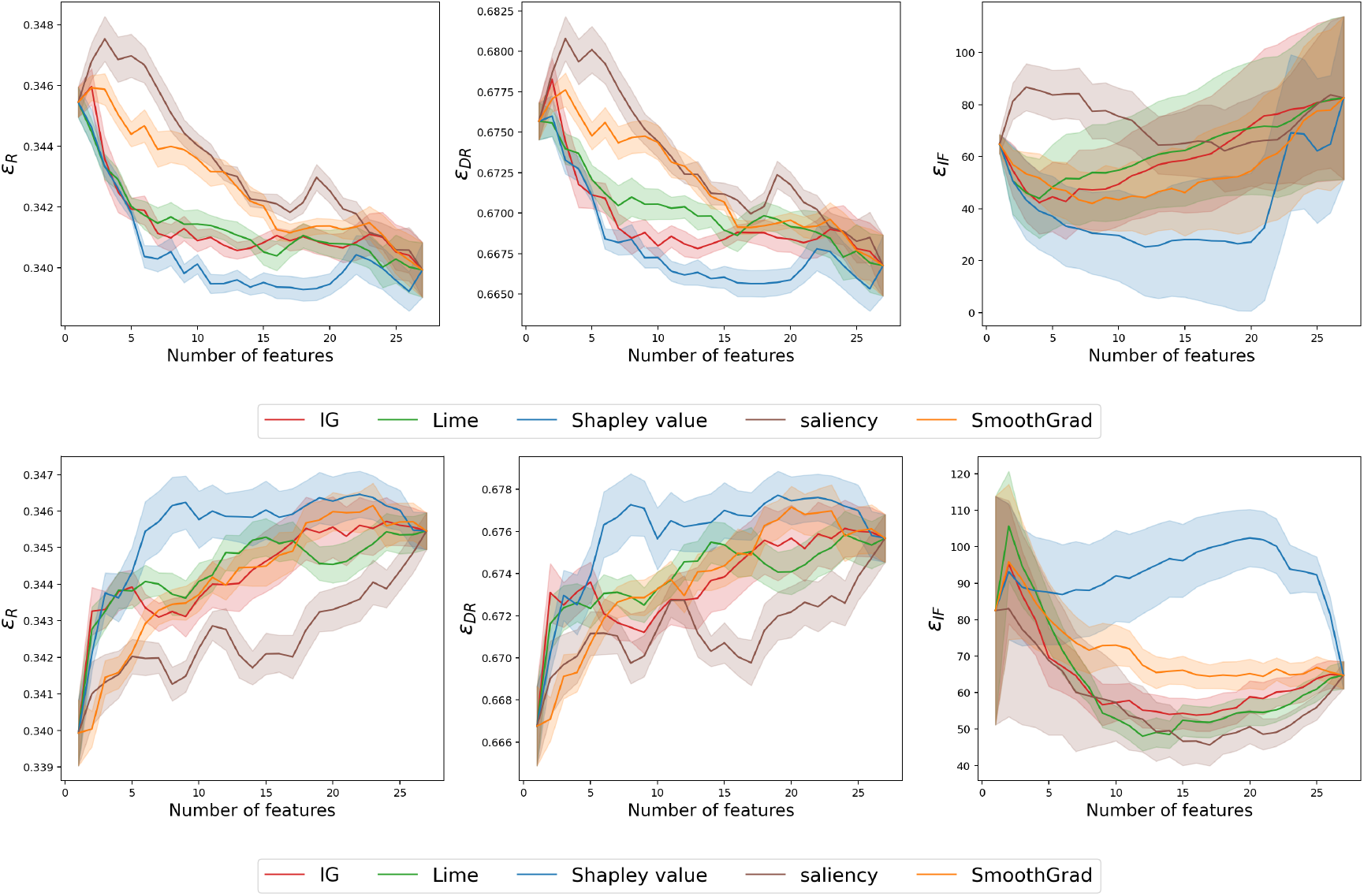
Insertion (top) and deletion (bottom) curves on the ACCORD dataset. The X-axis denotes the number of features removed or included, while the Y-axis shows the pseudo-outcome surrogates ℰ_*R*_, ℰ_*DR*_, and ℰ_*IF*_ .

**Fig. S.2.**
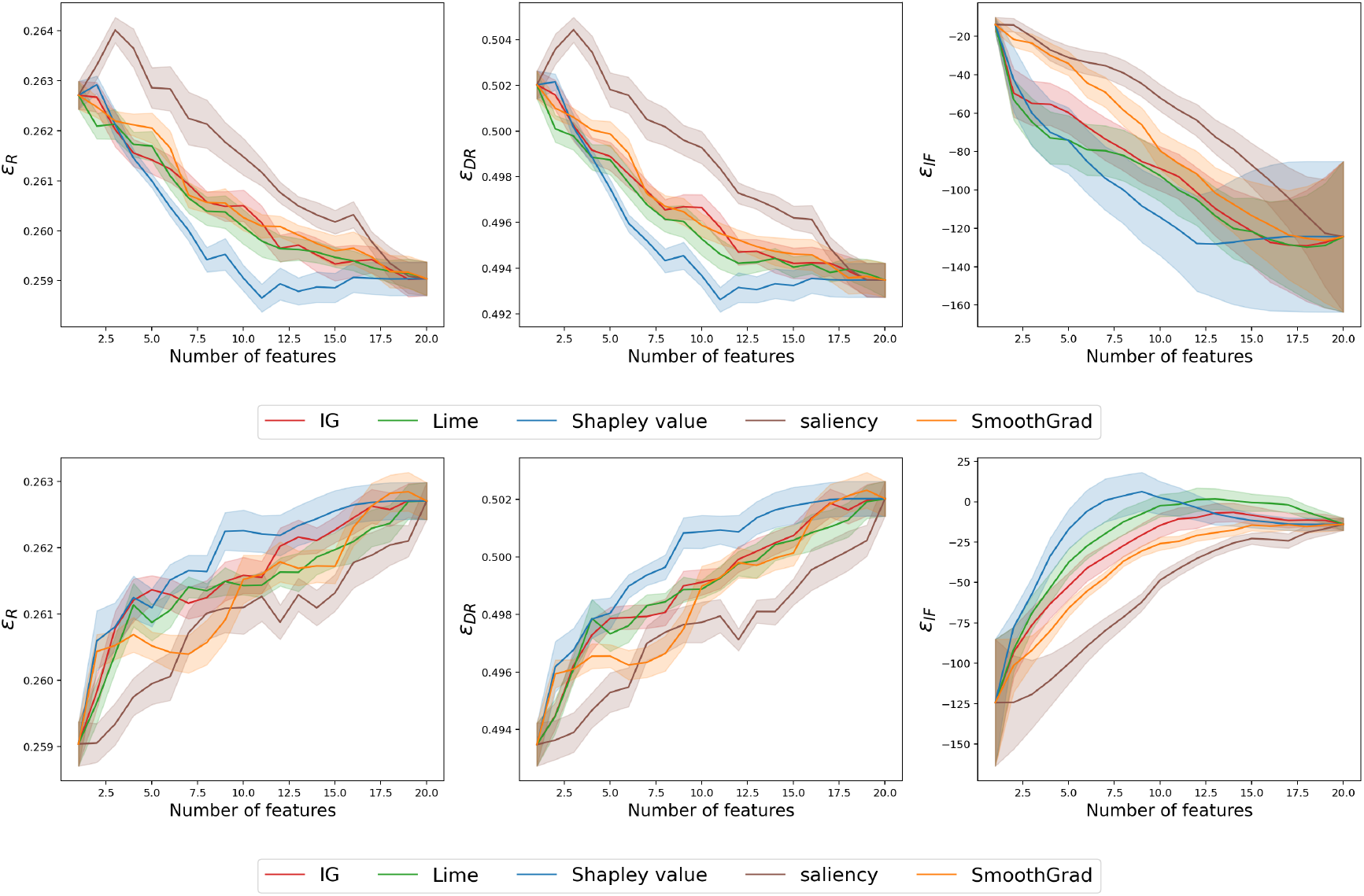
Insertion (top) and deletion (bottom) curves on the SPRINT dataset. The X-axis denotes the number of features removed or included, while the Y-axis shows the pseudo-outcome surrogates ℰ_*R*_, ℰ_*DR*_, and ℰ_*IF*_ .

**Fig. S.3.**
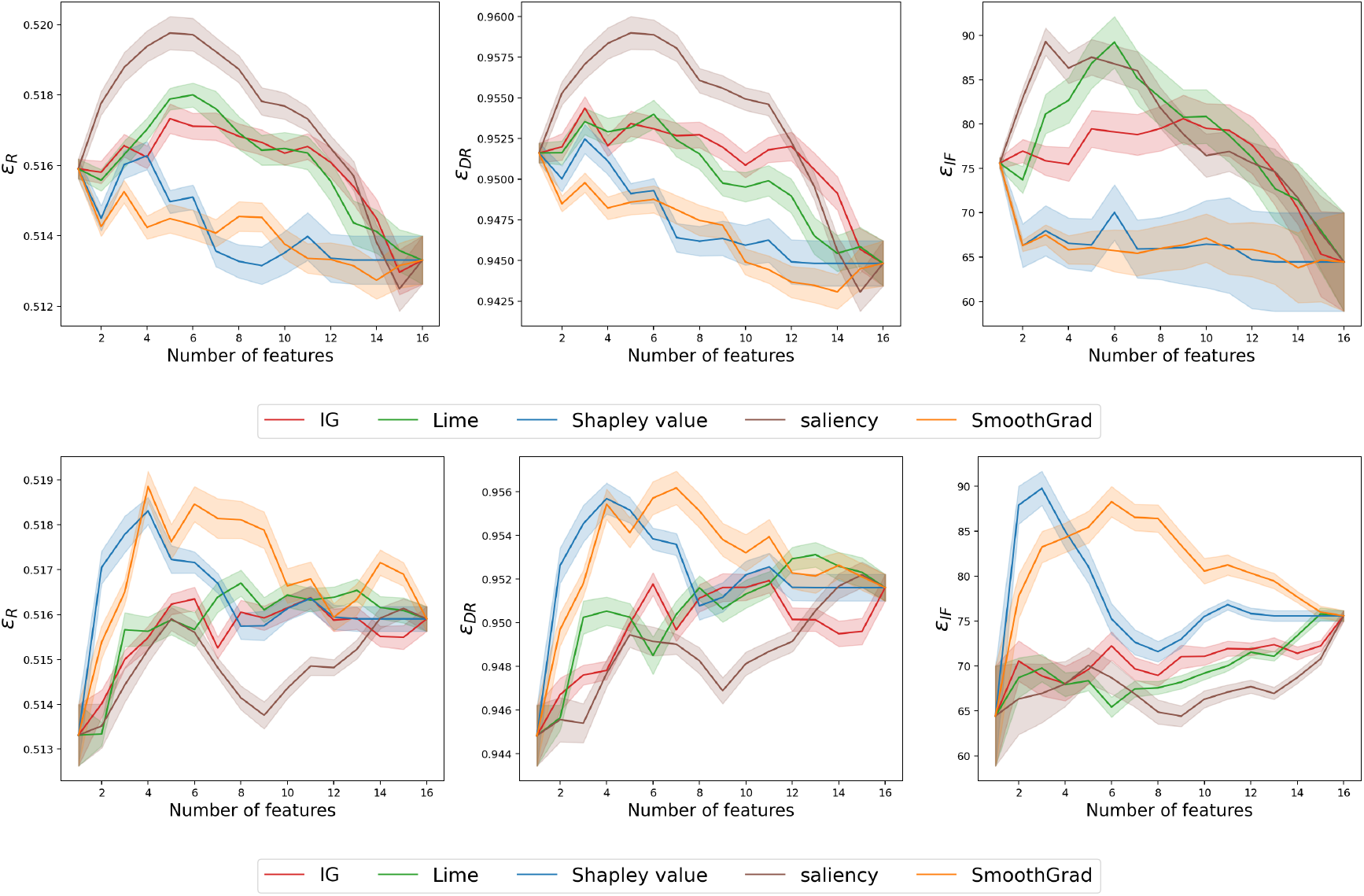
Insertion (top) and deletion (bottom) curves on the IST-3 dataset. The X-axis denotes the number of features removed or included, while the Y-axis shows the pseudo-outcome surrogates ℰ_*R*_, ℰ_*DR*_, and ℰ_*IF*_ .

**Fig. S.4.**
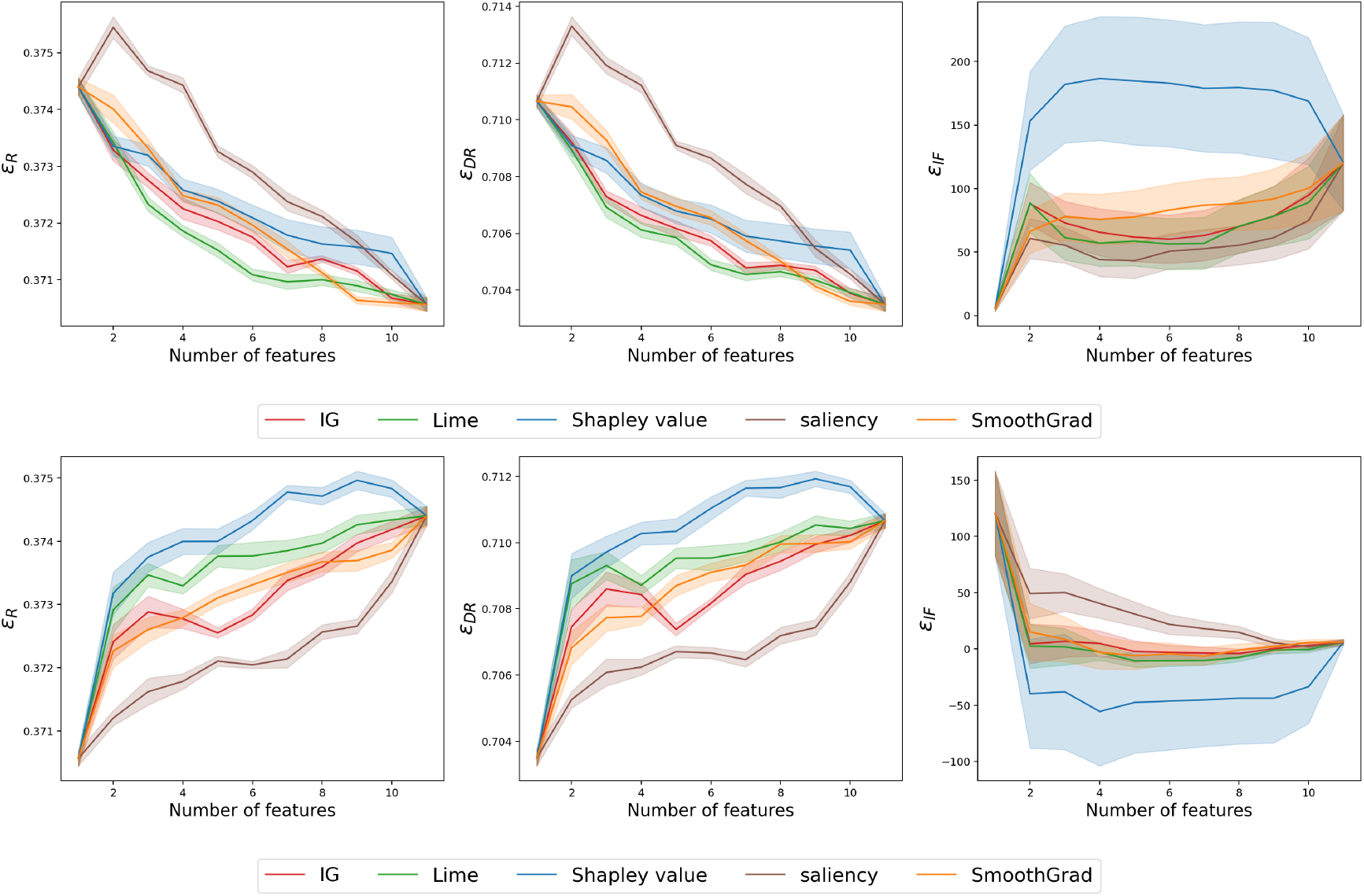
Insertion (top) and deletion (bottom) curves on the CRASH-2 dataset. The X-axis denotes the number of features removed or included, while the Y-axis shows the pseudo-outcome surrogates ℰ_*R*_, ℰ_*DR*_, and ℰ_*IF*_ .

**Fig. S.5.**
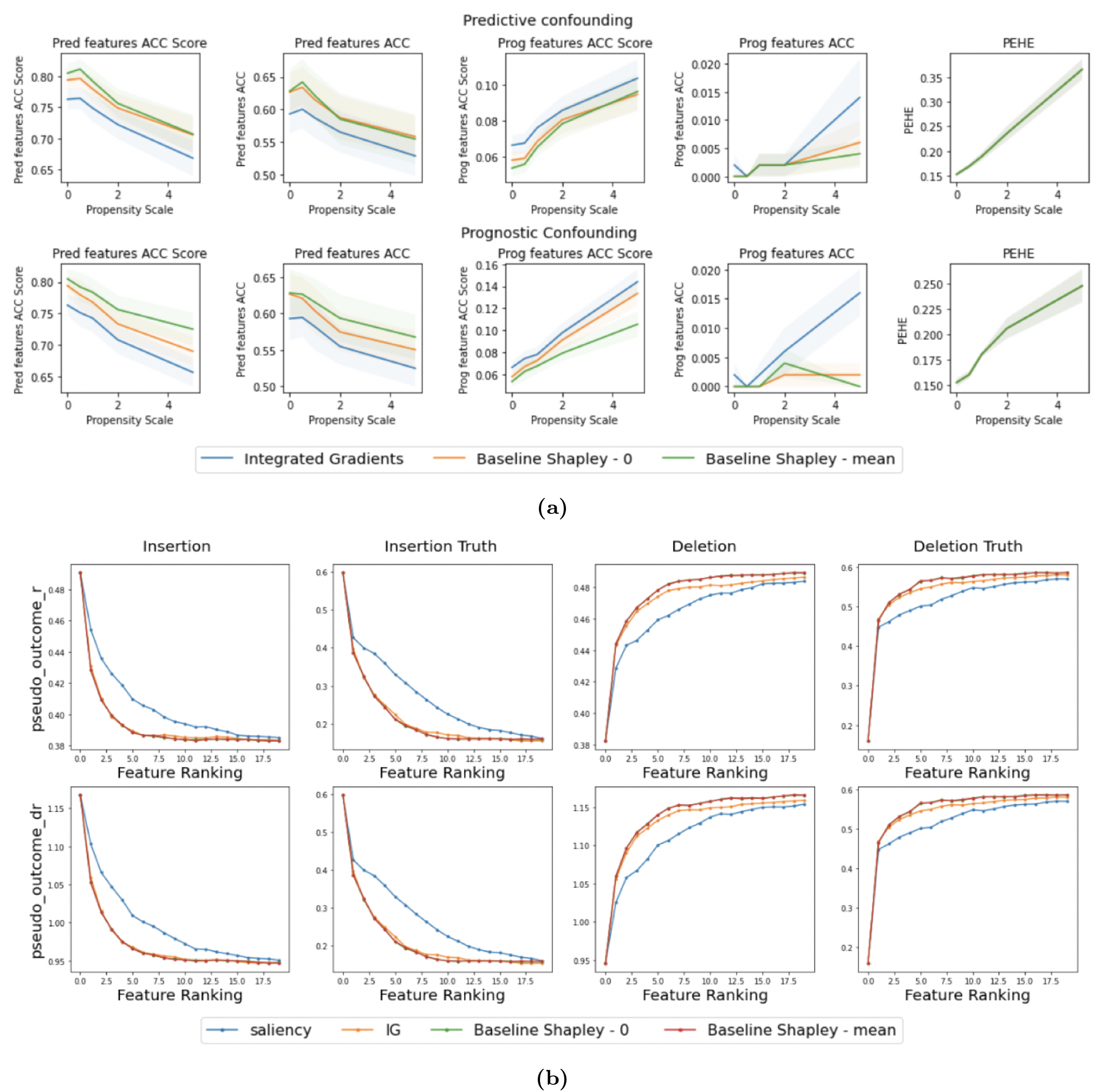
**(a)** Identifying Predictive & Prognostic Features in confounding environment with semisynthetic data: ACIC dataset comprises 20 predictive (10 pred_0_ + 10 pred_1_) and 10 prognostic features among a total of 55. The x-axis represents the (confounding) propensity scale, *ω*, while the y-axis depicts the percentage of features identified. **(b)Analyzing Insertion and Deletion via Pseudo-outcome surrogates**: This illustrates insertion (left panel) and deletion (right panel) analyses, employing both the R-Learner objective, 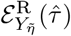 (top), and the DR-Learner objective, 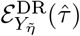 (bottom). The ground truth, *ϵ*, in the ACIC dataset is used with a propensity scale of *ω* = 0.5.

**Fig. S.6.**
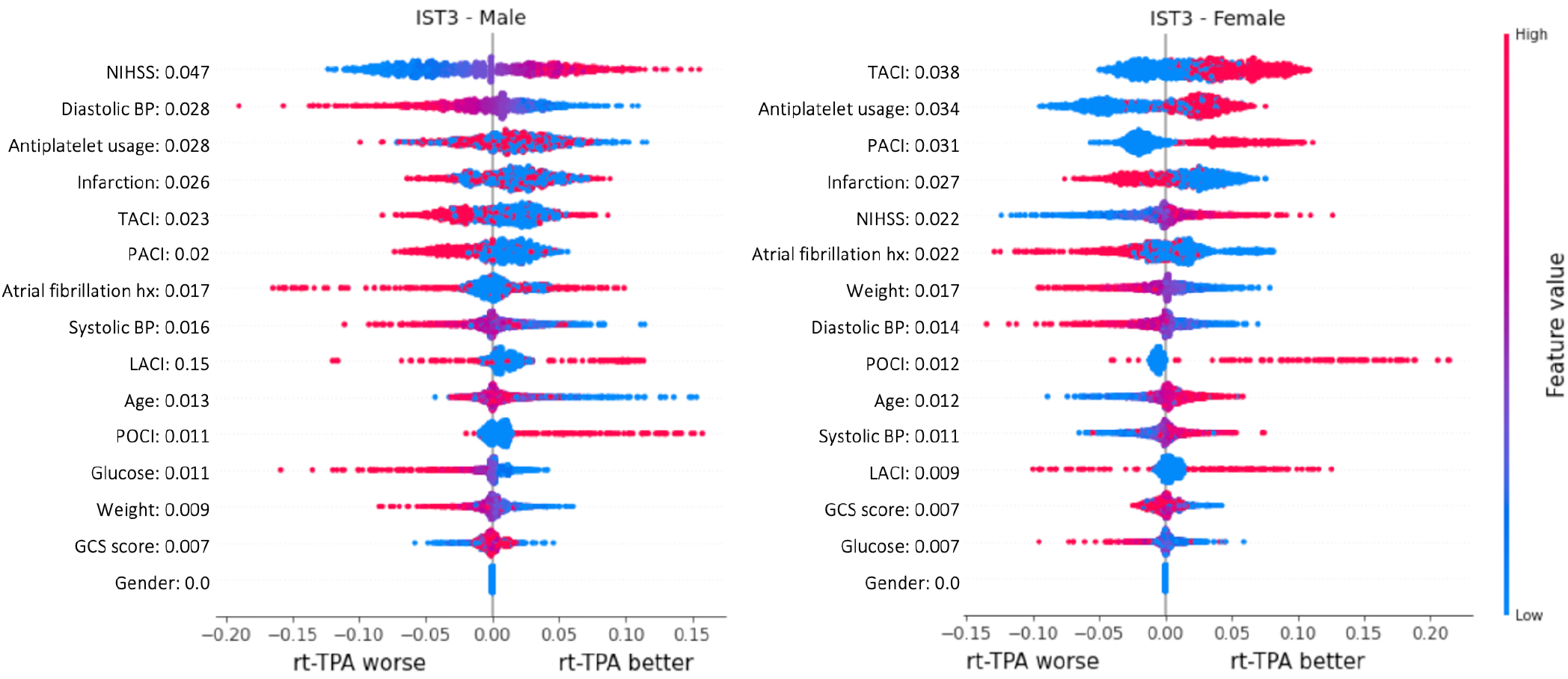
Shapley summary plots for the male (left) and female (right) subpopulations with gender-specific baselines.

**Fig. S.7.**
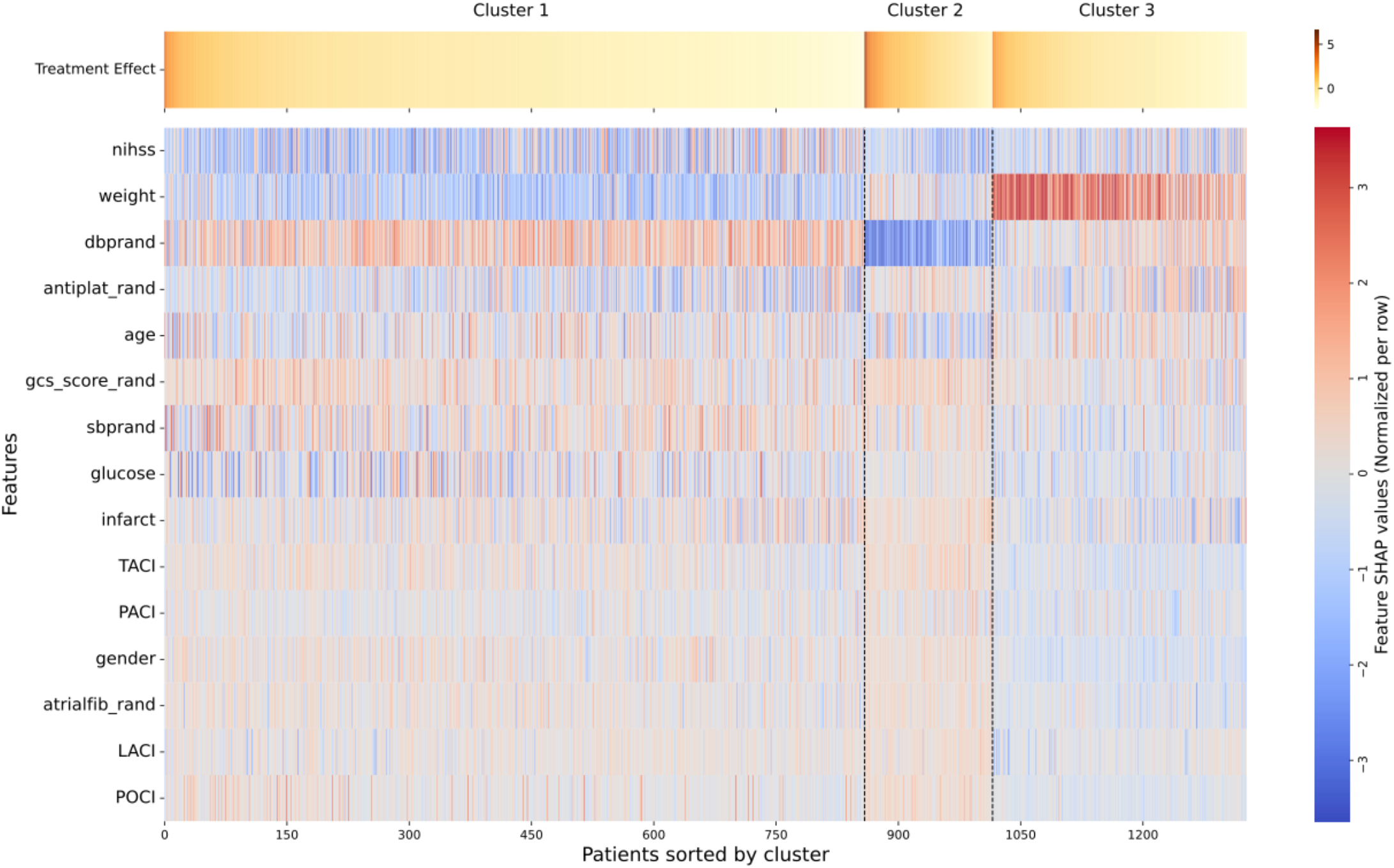
Hierarchical clustering of IST-3 patients with NIHSS scores between 6 and 14, based on Shapley value explanations.

**Fig. S.8.**
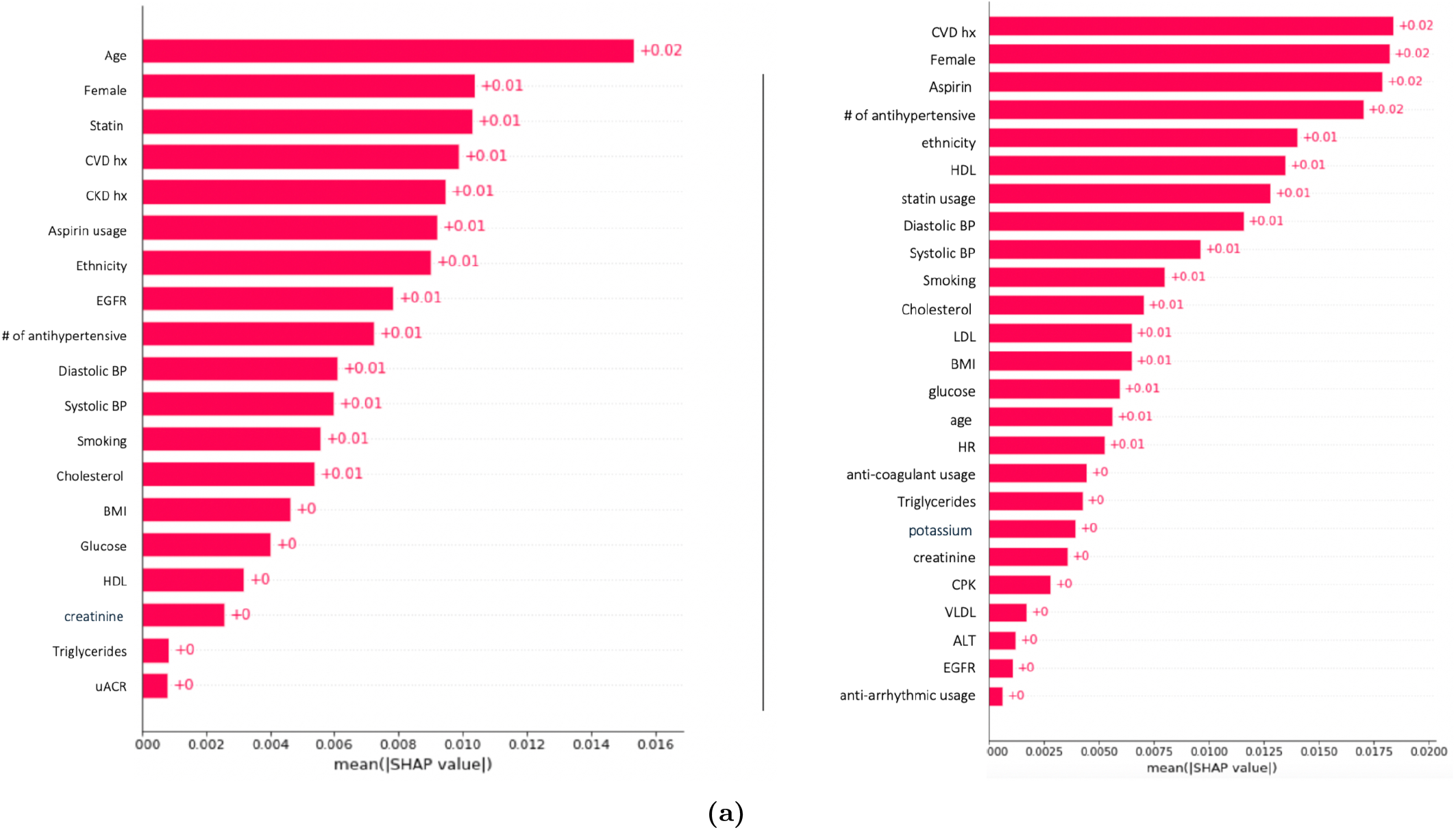
**(a)** Feature ranking between SPRINT (right) and ACCORD (left) based on absolute Shapley Values. The Y-axis represents feature names, and the X-axis indicates their average absolute Shapley values.

**Fig. S.9.**
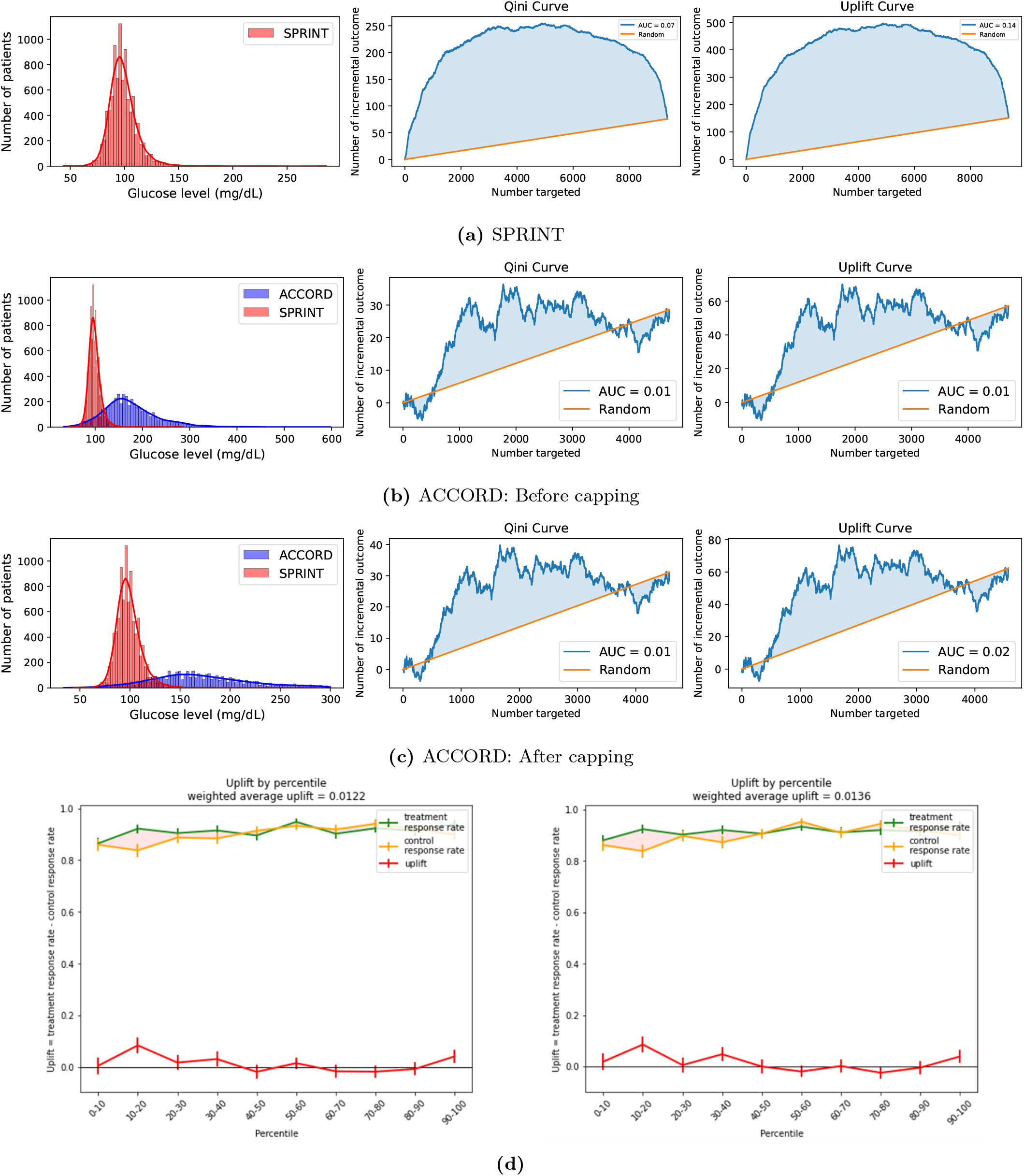
(a) Glucose distribution, Qini curve, and uplift curve for SPRINT. (b) Distribution of glucose levels, Qini curve, and uplift curve before capping at the maximum SPRINT glucose threshold. (c) Distribution of glucose levels, Qini curve, and uplift curve after capping at the maximum SPRINT glucose threshold. (d) Weighted uplift score, treatment response rate, and control response rate at each percentile before (left) and after (right) capping.

**Fig. S.10.**
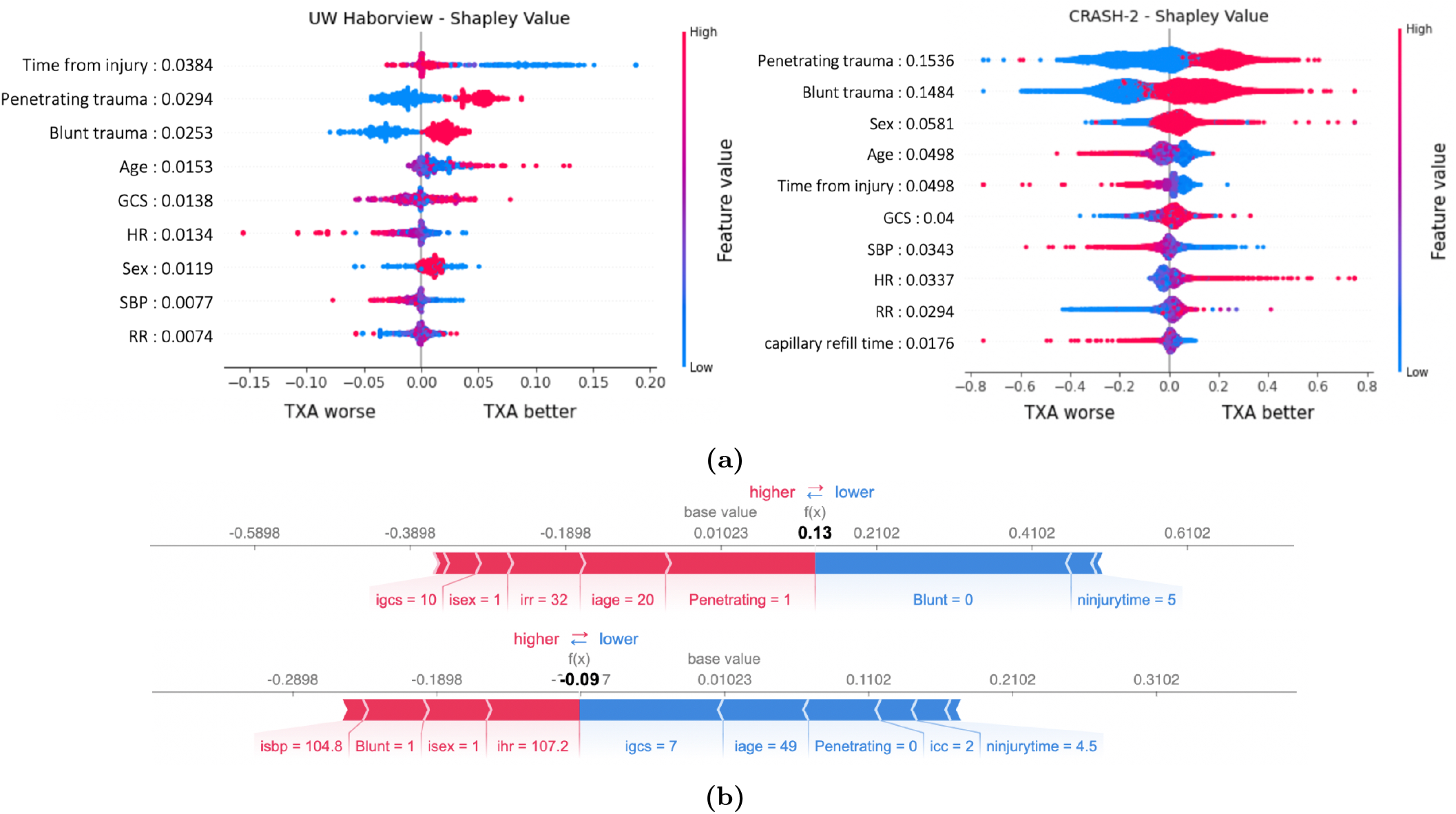
**(a)** Shapley summary plot for the Harborview cohort (left), CRASH-2 (right).**(b)** CRASH-2: Explaining sample individuals with different demographics and laboratory values. Red colors denote positive attributions and blue denotes negative attributions.

**Fig. S.11.**
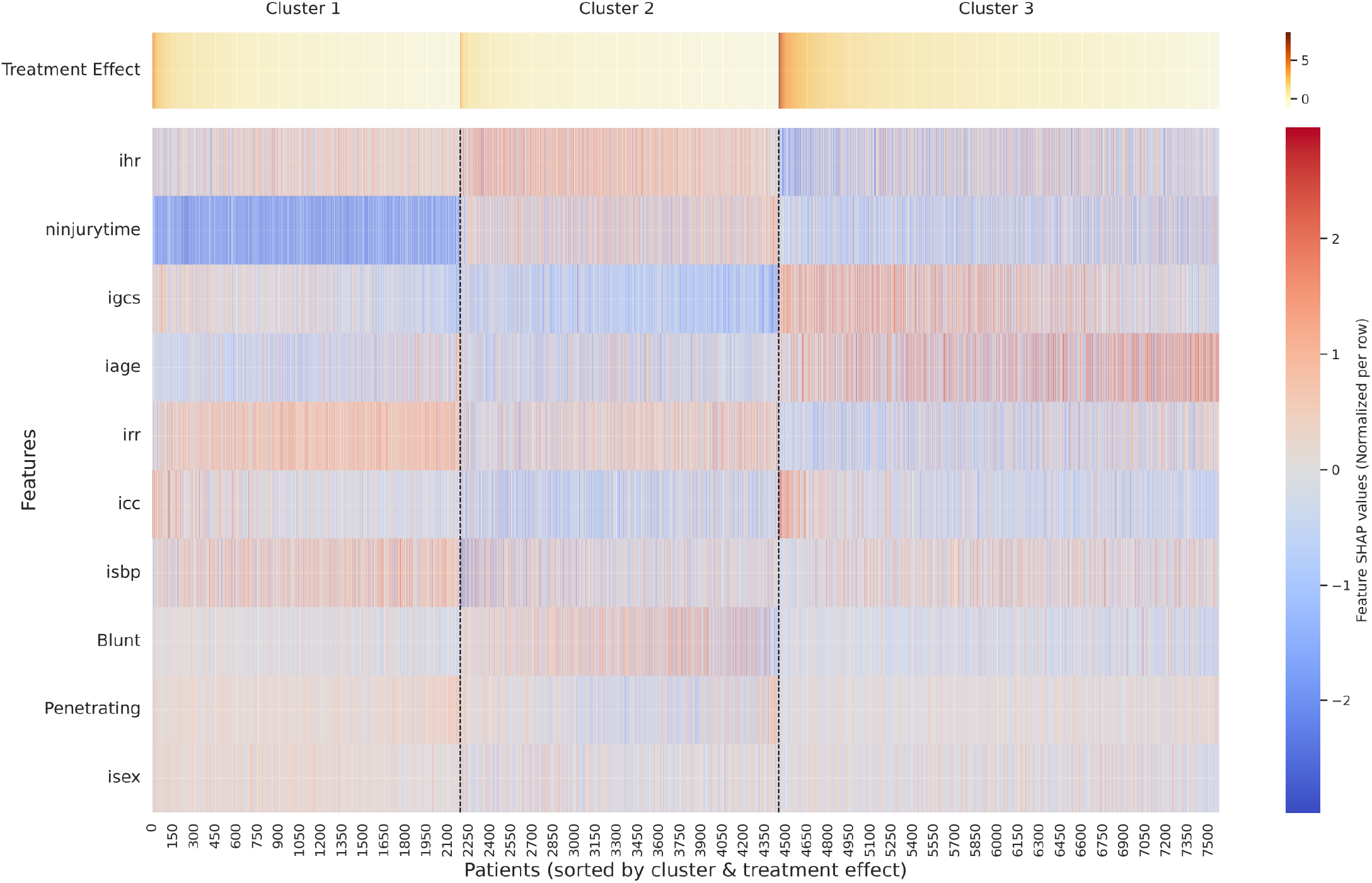
Hierarchical clustering of CRASH-2 patients with NIHSS scores between 6 and 14, based on Shapley value explanations.

**Table S.1.** Hyperparameter ranges for CATE models.

| Hyperparameter | Range |
| --- | --- |
| Training epochs | {10, 20, 30, 40, 50, 80, 100} |
| Learning rate | {$10^{-2}$, $10^{-3}$, $10^{-4}$, $10^{-5}$} |
| Batch size | {32, 64, 128} |
| Number of hidden layers | {1, 2, 4} |
| Hidden dimensions | {128, 256, 512} |
| Optimizer | Adam |
| Regularization constant (CFR) | {0, $10^{-1}$, $10^{-2}$, $10^{-3}$} |

**Table S.2.** Model performance on IST-3, CRASH-2, SPRINT, and ACCORD. Metrics include pseudo-surrogate outcomes ( ℰ_if-pehe_, ℰ_R_, ℰ_DR_) and incremental gain scores (Qini and Uplift). Results are reported as mean *±* standard deviation across random seeds. Higher values indicate better performance for Qini and Uplift, whereas lower values indicate better performance for ℰ_if-pehe_, ℰ_R_, and ℰ_DR_.

| Metric | XLearner | DRLearner | SLearner | RALearner | RLearner | TLearner | TARNet | DragonNet | CFRNet | CausalForest | LinearDR |
| --- | --- | --- | --- | --- | --- | --- | --- | --- | --- | --- | --- |
| IST-3 |  |  |  |  |  |  |  |  |  |  |  |
| Qini | 0.191 ± 0.023 | **0.284 ± 0.034** | 0.108 ± 0.023 | 0.182 ± 0.020 | 0.149 ± 0.017 | 0.169 ± 0.023 | 0.009 ± 0.019 | 0.009 ± 0.026 | -0.001 ± 0.013 | 0.214 ± 0.008 | 0.043 ± 0.013 |
| Uplift | 0.203 ± 0.025 | **0.300 ± 0.037** | 0.116 ± 0.024 | 0.193 ± 0.020 | 0.157 ± 0.017 | 0.181 ± 0.024 | 0.009 ± 0.020 | 0.009 ± 0.028 | -0.001 ± 0.014 | 0.233 ± 0.011 | 0.045 ± 0.013 |
| $\epsilon_{\text{if-pehe}}$ | 154.7 ± 127.7 | 250.0 ± 136.0 | 101.0 ± 128.4 | 138.6 ± 120.7 | 168.7 ± 129.1 | 129.9 ± 134.2 | 94.1 ± 97.6 | 94.6 ± 95.6 | 93.1 ± 96.2 | **73.9 ± 112.4** | 88.1 ± 108.8 |
| $\epsilon_R$ | 0.523 ± 0.006 | **0.521 ± 0.008** | 0.531 ± 0.007 | 0.526 ± 0.007 | 0.527 ± 0.008 | 0.526 ± 0.007 | 0.534 ± 0.007 | 0.534 ± 0.008 | 0.534 ± 0.008 | 0.526 ± 0.008 | 0.533 ± 0.007 |
| $\epsilon_{\text{DR}}$ | 0.924 ± 0.012 | **0.921 ± 0.016** | 0.941 ± 0.014 | 0.929 ± 0.013 | 0.932 ± 0.014 | 0.931 ± 0.013 | 0.947 ± 0.013 | 0.947 ± 0.013 | 0.947 ± 0.013 | 0.930 ± 0.014 | 0.946 ± 0.013 |
| ACCORD |  |  |  |  |  |  |  |  |  |  |  |
| Qini | 0.088 ± 0.006 | 0.105 ± 0.007 | 0.056 ± 0.006 | **0.147 ± 0.006** | 0.078 ± 0.008 | 0.097 ± 0.008 | -0.002 ± 0.008 | 0.003 ± 0.009 | -0.001 ± 0.008 | 0.100 ± 0.007 | 0.025 ± 0.004 |
| Uplift | 0.166 ± 0.011 | 0.191 ± 0.012 | 0.106 ± 0.012 | 0.187 ± 0.013 | 0.148 ± 0.014 | 0.184 ± 0.015 | -0.003 ± 0.014 | 0.005 ± 0.016 | -0.002 ± 0.016 | **0.272 ± 0.009** | 0.049 ± 0.008 |
| $\epsilon_{\text{if-pehe}}$ | 34.0 ± 74.4 | 124.5 ± 69.0 | -24.1 ± 66.8 | -7.3 ± 67.6 | 52.9 ± 65.9 | 1.3 ± 66.1 | 23.2 ± 42.8 | 21.4 ± 45.5 | 23.3 ± 44.3 | **-68.9 ± 81.3** | -10.8 ± 60.5 |
| $\epsilon_R$ | 0.310 ± 0.005 | 0.307 ± 0.006 | 0.318 ± 0.005 | **0.305 ± 0.006** | 0.311 ± 0.007 | 0.307 ± 0.007 | 0.320 ± 0.006 | 0.320 ± 0.006 | 0.320 ± 0.006 | 0.313 ± 0.006 | 0.320 ± 0.006 |
| $\epsilon_{\text{DR}}$ | 0.601 ± 0.010 | 0.595 ± 0.011 | 0.618 ± 0.010 | **0.592 ± 0.012** | 0.603 ± 0.013 | 0.596 ± 0.012 | 0.624 ± 0.009 | 0.623 ± 0.010 | 0.623 ± 0.010 | 0.609 ± 0.010 | 0.622 ± 0.010 |
| SPRINT |  |  |  |  |  |  |  |  |  |  |  |
| Qini | 0.059 ± 0.005 | 0.066 ± 0.003 | 0.035 ± 0.004 | 0.056 ± 0.004 | 0.054 ± 0.005 | 0.064 ± 0.002 | -0.000 ± 0.006 | 0.001 ± 0.004 | 0.003 ± 0.004 | **0.101 ± 0.006** | 0.014 ± 0.002 |
| Uplift | 0.114 ± 0.009 | 0.125 ± 0.005 | 0.068 ± 0.007 | 0.107 ± 0.007 | 0.104 ± 0.010 | 0.124 ± 0.004 | -0.001 ± 0.011 | 0.001 ± 0.007 | 0.006 ± 0.008 | **0.192 ± 0.009** | 0.026 ± 0.004 |
| $\epsilon_{\text{if-pehe}}$ | -82.9 ± 84.3 | -26.0 ± 81.5 | -111.9 ± 91.4 | -107.4 ± 87.4 | -43.5 ± 95.3 | -83.3 ± 81.9 | 37.5 ± 59.8 | 36.1 ± 61.4 | 34.0 ± 59.8 | **-157.7 ± 89.6** | -59.8 ± 68.9 |
| $\epsilon_R$ | 0.243 ± 0.005 | **0.241 ± 0.004** | 0.249 ± 0.005 | 0.244 ± 0.005 | 0.244 ± 0.005 | 0.242 ± 0.005 | 0.251 ± 0.005 | 0.251 ± 0.005 | 0.251 ± 0.005 | 0.245 ± 0.005 | 0.250 ± 0.005 |
| $\epsilon_{\text{DR}}$ | 0.476 ± 0.007 | **0.473 ± 0.007** | 0.488 ± 0.008 | 0.478 ± 0.008 | 0.477 ± 0.008 | 0.474 ± 0.008 | 0.492 ± 0.008 | 0.492 ± 0.008 | 0.492 ± 0.008 | 0.479 ± 0.008 | 0.491 ± 0.008 |
| CRASH-2 |  |  |  |  |  |  |  |  |  |  |  |
| Qini | 0.067 ± 0.004 | **0.136 ± 0.004** | 0.041 ± 0.004 | 0.054 ± 0.004 | 0.053 ± 0.005 | 0.066 ± 0.005 | 0.001 ± 0.005 | -0.000 ± 0.004 | 0.004 ± 0.004 | 0.072 ± 0.007 | 0.011 ± 0.003 |
| Uplift | 0.124 ± 0.009 | **0.248 ± 0.005** | 0.076 ± 0.007 | 0.099 ± 0.007 | 0.098 ± 0.009 | 0.122 ± 0.009 | 0.003 ± 0.009 | -0.000 ± 0.008 | 0.008 ± 0.007 | 0.132 ± 0.013 | 0.020 ± 0.005 |
| $\epsilon_{\text{if-pehe}}$ | -42.9 ± 177.9 | 139.8 ± 213.9 | -85.5 ± 165.3 | -60.5 ± 186.6 | 27.1 ± 191.8 | -12.9 ± 162.1 | 87.9 ± 131.2 | 93.0 ± 128.1 | 91.4 ± 131.7 | **-122.6 ± 172.4** | -8.4 ± 149.6 |
| $\epsilon_R$ | 0.374 ± 0.003 | **0.373 ± 0.003** | 0.377 ± 0.003 | 0.375 ± 0.003 | 0.376 ± 0.003 | 0.374 ± 0.003 | 0.379 ± 0.003 | 0.379 ± 0.003 | 0.379 ± 0.003 | 0.372 ± 0.003 | 0.379 ± 0.003 |
| $\epsilon_{\text{DR}}$ | 0.686 ± 0.005 | **0.684 ± 0.005** | 0.694 ± 0.004 | 0.690 ± 0.005 | 0.690 ± 0.005 | 0.687 ± 0.004 | 0.698 ± 0.004 | 0.697 ± 0.004 | 0.697 ± 0.004 | 0.683 ± 0.004 | 0.697 ± 0.004 |

**Table S.3.** Comparison of predicted average treatment effect (ATE) estimates from CATE models (with 95% confidence intervals) and reported ATE values (primary outcome differences) across four clinical trials.

| Cohort | Predicted ATE (95% CI) | Reported ATE (%) |
| --- | --- | --- |
| CRASH-2 | 1.1 (0.2 - 1.9) | 1.5 |
| IST-3 | 2.0 (0.3 - 4.0) | 2.0 |
| SPRINT | 1.6 (0.8 - 2.4) | 0.54 |
| ACCORD | 1.2 (-0.3 - 2.4) | 0.22 |

**Table S.4.** Normalized Area Under the Curve (AUC) for insertion and deletion curves on the ACCORD dataset. All values are normalized by the number of features. Results are reported for three pseudo-outcome surrogates: ℰ_*R*_, ℰ_*DR*_, and ℰ_*IF*_ . The best-performing method for each row is shown in **bold**.

| Metric | Shapley value | IG | Lime | Saliency | SmoothGrad |
| --- | --- | --- | --- | --- | --- |
| **Insertion** |  |  |  |  |  |
| $\mathcal{E}_{DR}$ (Del) ($\times 10^{-3}$) | **3.447** | 2.197 | 1.163 | 2.060 | 0.926 |
| $\mathcal{E}_R$ (Del) ($\times 10^{-3}$) | **2.314** | 1.462 | 1.357 | 0.854 | 0.341 |
| $\mathcal{E}_{IF}$ (Del) | **34.505** | 13.144 | 11.143 | 9.820 | 19.497 |
| **Deletion** |  |  |  |  |  |
| $\mathcal{E}_{DR}$ (Ins) [$\times 10^{-3}$] | **4.748** | 2.586 | 2.734 | 1.508 | 2.579 |
| $\mathcal{E}_R$ (Ins) [$\times 10^{-3}$] | **2.797** | 1.702 | 1.743 | 0.707 | 1.489 |
| $\mathcal{E}_{IF}$ (Ins) | **19.214** | 12.245 | 15.049 | 16.517 | 4.833 |

**Table S.5.** Normalized Area Under the Curve (AUC) for insertion and deletion curves on the SPRINT dataset. All values are normalized by the number of features. Results are reported for three pseudo-outcome surrogates: ℰ, ℰ_*DR*_, and ℰ_*IF*_ . The best-performing method for each row is shown in **bold**.

| Metric | Shapley value | IG | Lime | Saliency | SmoothGrad |
| --- | --- | --- | --- | --- | --- |
| **Insertion** |  |  |  |  |  |
| $\mathcal{E}_{DR}$ (Del) ($\times 10^{-3}$) | **2.573** | 1.294 | 1.725 | 1.079 | 1.140 |
| $\mathcal{E}_R$ (Del) ($\times 10^{-3}$) | **1.066** | 0.485 | 0.547 | 0.551 | 0.340 |
| $\mathcal{E}_{IF}$ (Del) | **33.271** | 22.262 | 27.084 | 8.440 | 8.914 |
| **Deletion** |  |  |  |  |  |
| $\mathcal{E}_{DR}$ (Ins) ($\times 10^{-3}$) | **2.289** | 1.356 | 1.228 | 0.591 | 0.937 |
| $\mathcal{E}_R$ (Ins) ($\times 10^{-3}$) | **1.044** | 0.776 | 0.583 | 0.272 | 0.497 |
| $\mathcal{E}_{IF}$ (Ins) | **51.544** | 38.708 | 47.688 | 12.237 | 29.173 |

**Table S.6.** Normalized Area Under the Curve (AUC) for insertion and deletion curves on the IST-3 dataset. Value is normalized by the number of features. Results are reported for three pseudo-outcome surrogates: ℰ_*R*_, ℰ_*DR*_, and ℰ_*IF*_ . The best-performing method for each row is shown in **bold**.

| Metric | Shapley value | IG | Lime | Saliency | SmoothGrad |
| --- | --- | --- | --- | --- | --- |
| **Insertion** |  |  |  |  |  |
| $\mathcal{E}_{DR}$ (Del) ($\times 10^{-3}$) | 0.833 | -3.156 | -2.005 | -5.552 | **1.625** |
| $\mathcal{E}_R$ (Del) ($\times 10^{-3}$) | 0.455 | -1.398 | -1.448 | -2.680 | **0.619** |
| $\mathcal{E}_{IF}$ (Del) | **3.621** | -6.141 | -8.672 | -9.476 | 3.902 |
| **Deletion** |  |  |  |  |  |
| $\mathcal{E}_{DR}$ (Ins) ($\times 10^{-3}$) | 4.237 | 1.611 | 2.453 | -0.448 | **4.921** |
| $\mathcal{E}_R$ (Ins) ($\times 10^{-3}$) | 1.825 | 1.009 | 1.253 | -0.279 | **2.431** |
| $\mathcal{E}_{IF}$ (Ins) | 7.401 | 0.649 | 3.831 | -2.425 | **11.364** |

**Table S.7.** Normalized Area Under the Curve (AUC) for insertion and deletion curves on the CRASH-2 dataset. All values are normalized by the number of features. Results are reported for three pseudo-outcome surrogates: ℰ_*R*_, ℰ_*DR*_, and ℰ_*IF*_ . The best-performing method for each row is shown in **bold**.

| Metric | Shapley value | IG | Lime | Saliency | SmoothGrad |
| --- | --- | --- | --- | --- | --- |
| **Insertion** |  |  |  |  |  |
| $\mathcal{E}_{DR}$ (Del) [$\times 10^{-3}$] | 0.281 | 1.042 | **1.351** | 1.522 | 0.450 |
| $\mathcal{E}_R$ (Del) [$\times 10^{-3}$] | 0.228 | 0.584 | **0.853** | 0.562 | 0.435 |
| $\mathcal{E}_{IF}$ (Del) | **102.527** | 8.601 | 4.775 | 7.077 | 17.944 |
| **Deletion** |  |  |  |  |  |
| $\mathcal{E}_{DR}$ (Ins) [$\times 10^{-3}$] | **3.357** | 1.495 | 2.282 | 0.286 | 1.568 |
| $\mathcal{E}_R$ (Ins) [$\times 10^{-3}$] | **1.618** | 0.623 | 1.127 | 0.288 | 0.645 |
| $\mathcal{E}_{IF}$ (Ins) | **96.215** | 56.202 | 60.736 | 33.583 | 55.773 |

**Table S.8.** Knowledge distillation performance and top 5 identified features for datasets IST-3, CRASH-2, SPRINT, and ACCORD. Black indicates the method with the lowest distillation loss with feature budgets at 5.

| Explanation Methods | MSE ($\times 10^{-3}$) | Top 5 Features |
| --- | --- | --- |
| CRASH-2 |  |  |
| Saliency | 4.0 ($\pm 0.2$) | heart rate, injury time, respiratory, capillary refill time, systolic blood pressure |
| SmoothGrad | 3.8 ($\pm 0.2$) | heart rate, injury time, respiratory, capillary refill time, systolic blood pressure |
| Shapley | 3.5 ($\pm 0.1$) | Injury type, gender, GCS, injury time, age |
| IG | 3.5 ($\pm 0.2$) | Injury type, gender, injury time, age |
| IST-3 |  |  |
| Saliency | 19.1 ($\pm 0.64$) | gcs, nihss, age, weight, infarct: possibly yes |
| SmoothGrad | 18.8 ($\pm 0.60$) | diastolic blood pressure, nihss, age, weight, gender |
| **Shapley** | **17.0 ($\pm 0.64$)** | **Nihss, infarct, anti-platelet usage, atrial fibrillation, gender** |
| IG | 17.0 ($\pm 0.66$) | Nihss, infarct, anti-platelet usage, atrial fibrillation, TACI |
| SPRINT |  |  |
| Saliency | 6.1 ($\pm 0.21$) | Umacr, EGFR, TRR, creatine, glucose |
| SmoothGrad | 5.9 ($\pm 0.21$) | creatinine, EGFR, age, diastolic blood pressure |
| Shapley | 5.1 ($\pm 0.12$) | CKD history, statin usage, Age, Cardiovascular history, gender |
| **IG** | **5.3 ($\pm 0.17$)** | **Age, gender, statin, CKD history, Cardiovascular history** |
| ACCORD |  |  |
| Saliency | 17.6 ($\pm 0.64$) | Trig, EGFR, Potassium, ALT, HDL |
| SmoothGrad | 17.0 ($\pm 0.64$) | HDL, potassium, EGFR, alt, Trig |
| **Shapley** | **14.7 ($\pm 0.56$)** | **Cardiovascular history, gender, aspirin, # of anti-bp agents, race** |
| IG | 14.8 ($\pm 0.57$) | Cardiovascular history, gender, aspirin, # of anti-bp agents, race |

**Table S.9.**
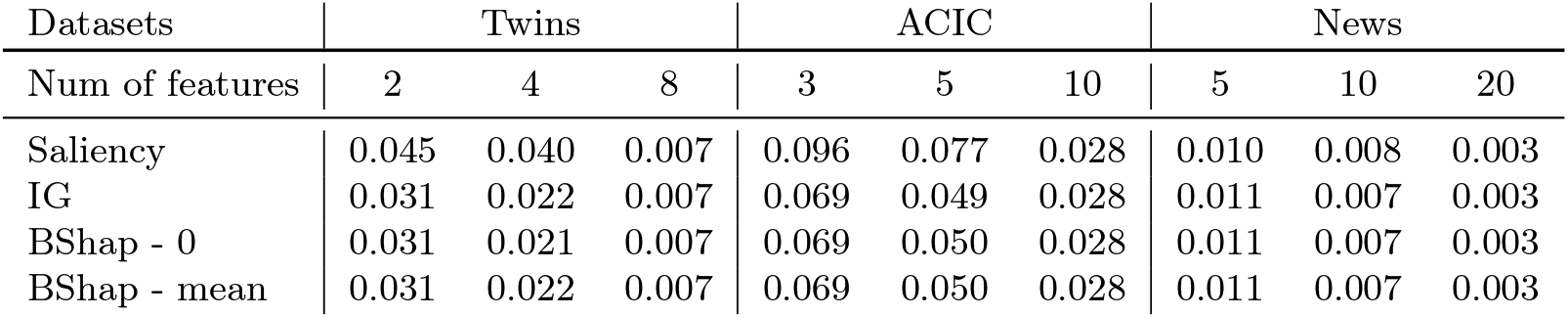
Knowledge Distillation Performance. Average MSE of student models relative to the original model, based on various feature counts in synthetic contexts. Environments encompass predictive and prognostic confounding with propensity scales *ω* = {0, 0.5, 1, 2, 5, 10}. Features are selected at rates of 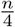 and 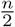, with *n* being the total feature count for semi-synthetic data: 8 (Twins), 10 (ACIC), and 20 (News).

**Table S.10.**
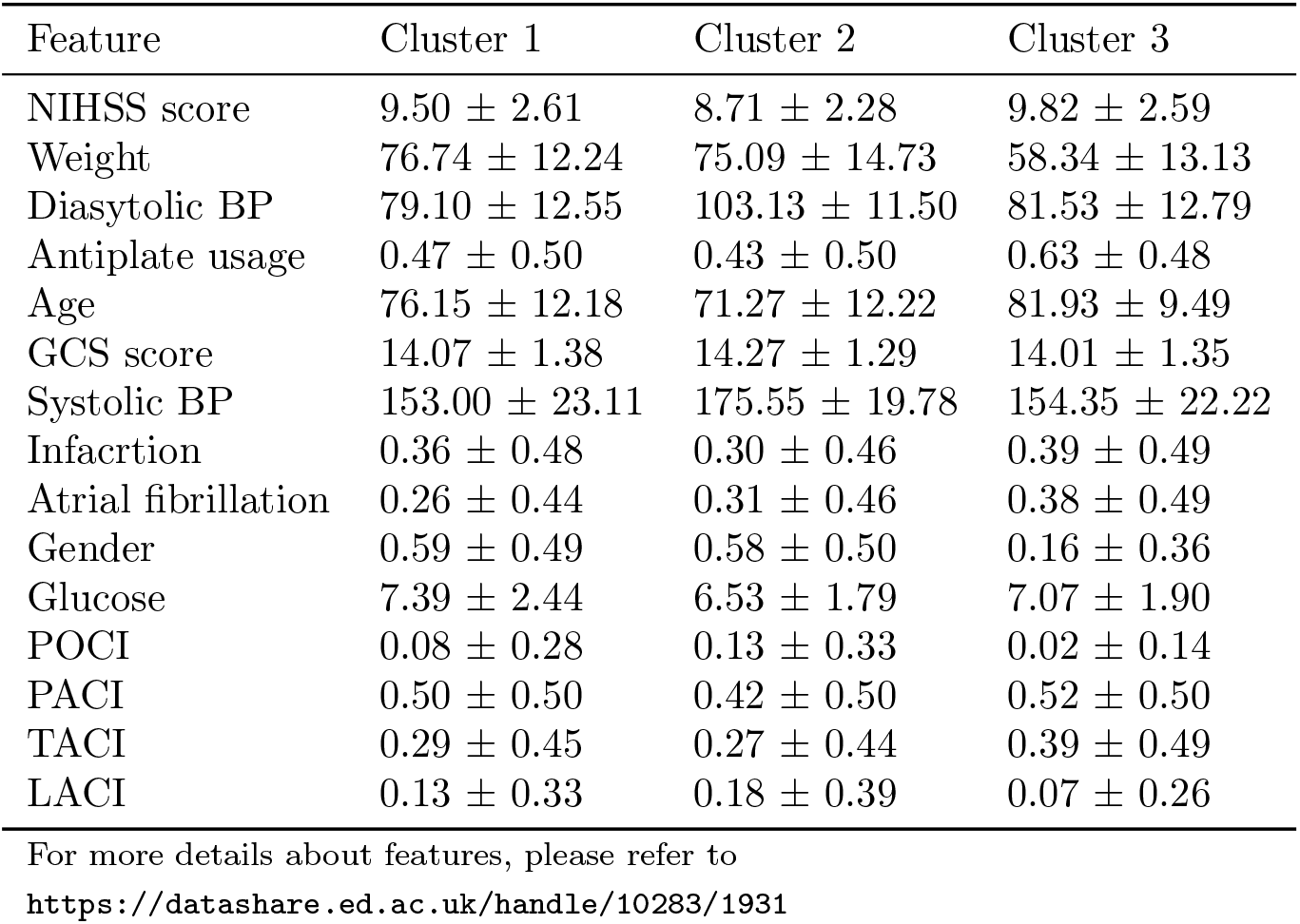
Cluster-wise feature summary (Mean *±* Std).

**Table S.11.** Uplift and Qini scores with 95% confidence intervals for various datasets. (*) denotes datasets excluding individuals with glucose levels greater than 300 mg/dL in ACCORD and patients older than 45 y/o in the Harborview trauma registry.

| Dataset | Uplift Score ($\times 10^{-2}$) | Qini Score ($\times 10^{-2}$) |
| --- | --- | --- |
| SPRINT (train) | $7.5 \pm 0.11$ | $3.9 \pm 0.06$ |
| ACCORD (test) | $0.38 \pm 0.17$ | $0.22 \pm 0.09$ |
| ACCORD* | $0.53 \pm 0.17$ | $0.30 \pm 0.09$ |
| CRASH-2 (train) | $7.6 \pm 0.11$ | $4.0 \pm 0.07$ |
| Harborview (test) | $0.05 \pm 0.04$ | $-0.05 \pm 0.03$ |
| Harborview* | $0.50 \pm 0.08$ | $0.08 \pm 0.05$ |

**Table S.12.** Cluster-wise summary of trauma features (Mean *±* Std).

| Feature | Cluster 1 | Cluster 2 | Cluster 3 |
| --- | --- | --- | --- |
| ninjurytime | 5.93 ± 1.43 | 3.98 ± 1.26 | 4.73 ± 1.51 |
| irr | 21.09 ± 4.79 | 22.47 ± 6.01 | 24.38 ± 7.76 |
| isbp | 107.67 ± 26.24 | 101.00 ± 23.91 | 100.61 ± 25.63 |
| ihr | 97.93 ± 19.15 | 102.29 ± 17.48 | 108.39 ± 24.09 |
| iage | 37.28 ± 11.96 | 33.14 ± 11.94 | 36.29 ± 17.80 |
| isex | 0.88 ± 0.33 | 0.83 ± 0.38 | 0.82 ± 0.38 |
| igcs | 12.68 ± 3.30 | 13.62 ± 2.50 | 10.66 ± 4.35 |
| icc | 3.45 ± 1.59 | 2.90 ± 1.44 | 3.48 ± 1.83 |
| Penetrating | 0.12 ± 0.33 | 0.48 ± 0.50 | 0.14 ± 0.35 |
| Blunt | 0.88 ± 0.33 | 0.52 ± 0.50 | 0.86 ± 0.35 |

\* isex (male = 1, female = 0) and type of trauma are encoded as binary variables. For more details on feature descriptions, please refer to the CRASH-2 data dictionary.

## Footnotes

1 https://github.com/AliciaCurth/CATENets

2 For local attribution methods, global rankings are obtained by averaging absolute attributions across individuals.

3 This procedure is conceptually similar to RemOve And Retrain (ROAR) ^48^, but evaluates explanations at the global level.

4 https://github.com/AliciaCurth/CATENets

5 https://github.com/jpaillard/permucate/blob/main/scripts/ihdp_exp.py

6 Note that this is different from the random treatment assignment in RCT.

7 https://captum.ai/

## Notes

### Competing Interest Statement

The authors have declared no competing interest.

### Funding Statement

This study did not receive any funding

### Author Declarations

The survey data for this study was gathered under an exempt determination from the University of Washington Institutional Review Board (Human Subjects Division, STUDY00006890). All necessary patient/participant consent has been obtained and the appropriate institutional forms have been archived.

### Summary of Updates

Update for corresponding authors' emails.

